# OpenSAFELY: The impact of COVID-19 on azathioprine, leflunomide, and methotrexate monitoring, and factors associated with change in monitoring rate

**DOI:** 10.1101/2023.06.06.23290826

**Authors:** The OpenSAFELY Collaborative, Andrew D. Brown, Louis Fisher, Helen J. Curtis, Milan Wiedemann, William J. Hulme, Lisa E.M. Hopcroft, Christine Cunningham, Victoria Speed, Ruth E. Costello, James B. Galloway, Mark D. Russell, Katie Bechman, Zeyneb Kurt, Richard Croker, Chris Wood, Alex J. Walker, Andrea L. Schaffer, Seb C.J. Bacon, Amir Mehrkar, George Hickman, Chris Bates, Jonathan Cockburn, John Parry, Frank Hester, Sam Harper, Ben Goldacre, Brian MacKenna

## Abstract

**Background:** The COVID-19 pandemic created unprecedented pressure on healthcare services. This study aimed to investigate if disease-modifying anti-rheumatic drug (DMARD) safety monitoring was affected during the COVID-19 pandemic.

**Methods:** A population-based cohort study was conducted with the approval of NHS England, using the OpenSAFELY platform to access electronic health record data from 24·2 million patients registered at general practices using TPP’s SystmOne software. Patients were included for further analysis if prescribed azathioprine, leflunomide, or methotrexate between November 2019 and July 2022. Outcomes were assessed as monthly trends and variation between various sociodemographic and clinical groups for adherence with standard safety monitoring recommendations.

**Findings:** An acute increase in the rate of missed monitoring occurred across the study population (+12·4 percentage points) when lockdown measures were implemented in March 2020. This increase was more pronounced for some patient groups (70-79 year-olds: +13·7 percentage points; females: +12·8 percentage points), regions (North West: +17·0 percentage points), medications (Leflunomide: +20·7 percentage points), and monitoring tests (Blood Pressure: +24·5 percentage points). Missed monitoring rates decreased substantially for all groups by July 2022. Substantial and consistent differences were observed in overall missed monitoring rates between several groups throughout the study.

**Interpretation:** DMARD monitoring rates temporarily deteriorated during the COVID-19 pandemic. Deterioration coincided with the onset of lockdown measures, with monitoring rates recovering rapidly as lockdown measures were eased. Differences observed in monitoring rates between medications, tests, regions, and patient groups, highlight opportunities to tackle potential inequalities in the provision or uptake of monitoring services. Further research should aim to evaluate the causes of the differences identified between groups.

**Funding:** None.

**Research in context:** *Evidence before this study:* Disease-modifying anti-rheumatic drugs (DMARDs) are immunosuppressive and/or immunomodulatory drugs, which carry risks of serious adverse effects such as; gastrointestinal, renal, hepatic, and pulmonary toxicity; myelosuppression; and increased susceptibility to infection. To mitigate these safety risks, national safety guidance recommends that patients taking these drugs receive regular monitoring. We searched PubMed, Web of Science and Scopus for studies published between database inception and July 28th, 2022, using the terms ([covid-19] AND [monitoring OR shared care OR dmard OR outcome factors] AND [primary care]), with no language restrictions. Studies that investigated the effect of the COVID-19 pandemic on healthcare services were identified. One key study in England showed disruption to various monitoring services in primary care had occurred during the pandemic. Another English study highlighted a disproportionate impact of the COVID-19 pandemic on health outcomes in certain groups.

*Added value of this study:* Prior to this study knowledge of how high-risk drugs, such as DMARDs, were affected by the COVID-19 pandemic was limited. This study reports the impact of COVID-19 on the safety monitoring of DMARDs. Moreover, it reports variation in DMARD monitoring rates between demographic, clinical and regional subgroups, which has not yet been described. This is enabled through use of the OpenSAFELY platform, which provides secure access to pseudonymised primary care patient records in England for the purposes of analysing the COVID-19 pandemic impact.

*Implications of all the available evidence:* DMARD monitoring rates transiently deteriorated during the COVID-19 pandemic, consistent with previous research on other monitoring tests. Deterioration coincided with the onset of lockdown measures, with performance recovering rapidly as lockdown measures were eased. Differences observed in monitoring rates between demographic, clinical and regional subgroups highlight opportunities to identify and tackle potential inequalities in the provision or uptake of monitoring services. Further research should aim to evaluate the causes of the differences identified between groups, and establish the clinical relevance of missed monitoring. Several studies have demonstrated the capability of the OpenSAFELY platform as a secure and efficient approach for analysing NHS primary care data at scale, generating meaningful insights on service delivery.

## Background

Disease-modifying anti-rheumatic drugs (DMARDs) are typically prescribed for autoimmune conditions such as rheumatoid arthritis, Crohn’s disease, and severe psoriasis. Many DMARDs have narrow therapeutic indexes, and can cause potentially fatal adverse events such as blood dyscrasias and liver toxicity.^1,2^ To mitigate these risks, DMARDs are typically initiated by a specialist clinician in secondary care. Once patients have been stabilised on treatment, they can be transferred to primary care under a shared care arrangement, which specifies long-term safety monitoring recommendations (Table 1) for General Practitioners (GPs) to oversee. Under the arrangement GPs have guidance on when to suspend a prescription or seek specialist advice, e.g. if they observe unsafe monitoring results, non-adherence to monitoring, or emergence of adverse effects.

**Table 1.** Examples of licensed indications typically seen in primary care, and long-term monitoring requirements for three commonly prescribed DMARDs.

| Drug Name | Indications | Long-term Monitoring |
| --- | --- | --- |
| Azathioprine | Crohn's disease, severe eczema, myasthenia gravis. | Every 12 weeks:<br>• Full Blood Count<br>• Urea & Electrolytes<br>• Liver Function Tests |
| Leflunomide | Rheumatoid arthritis, psoriatic arthritis. | Every 12 weeks:<br>• Full Blood Count<br>• Urea & Electrolytes<br>• Liver Function Tests<br>• Blood Pressure<br>• Weight |
| Methotrexate | Crohn's disease, severe psoriasis, rheumatoid arthritis. | Every 12 weeks:<br>• Full Blood Count<br>• Urea & Electrolytes<br>• Liver Function Tests |

The first wave of the COVID-19 pandemic in March 2020 prompted a national lockdown in the UK. GP Practices made several changes with the aim of reducing viral transmission, ranging from reducing face-to-face appointments, to temporary practice closure where outbreaks occurred amongst practice staff.^3^ Moreover, many patients worldwide chose to avoid healthcare services for fear of contracting COVID-19,^4,5,6^ and those who were recommended to isolate through ‘shielding’ faced additional barriers to healthcare access.^7^ In April 2020, the British Medical Association & Royal College of GP’s issued guidance suggesting clinicians consider DMARD monitoring a high priority task.^8^ However, The NHS Specialist Pharmacy Service and British Society of Rheumatology suggested that monitoring intervals for certain medications, including DMARDs, could be considered for extension.^9,10^ Several laboratory tests in GP practices experienced substantial activity reductions during lockdown,^11^ including tests related to methotrexate monitoring.^12^ However, previous work did not indicate whether specific patient groups were affected.

OpenSAFELY is a secure analytics platform for electronic patient records built with the approval of NHS England to deliver urgent academic^13^ and operational NHS service research^11,14^ on the impacts of the pandemic. Analyses run across patients’ full raw pseudonymised primary care records at English GP practices with patient-level linkage to sources of secondary care data.

We set out to assess the impact of COVID-19 on the monitoring of DMARDs, and evaluate whether any such effect was associated with other variables, such as patient demographics or comorbidities.

## Methods

### Study Design

We conducted a retrospective cohort study across 24 million patients by using EHR data from all GP practices in England supplied by software provider TPP. From this data we sought to identify patients prescribed DMARDs between November 2019 and July 2022.

### Data Source

Primary care records managed by TPP are available in OpenSAFELY, a data analytics platform created by our team on behalf of NHS England to address urgent COVID-19 research questions (https://opensafely.org). OpenSAFELY provides a secure software interface allowing analysis of pseudonymised primary care patient records from England in near real-time within the EHR vendor’s secure data centre, avoiding the need for large volumes of potentially disclosive pseudonymised patient data to be transferred off-site. This, in addition to other technical and organisational controls, minimises risk of re-identification. Similarly pseudonymised datasets from other data providers are securely provided to the EHR vendor and linked to the primary care data. The dataset analysed within OpenSAFELY is based on people currently registered with GP practices using TPP SystmOne software. It includes pseudonymised data such as coded diagnoses, medications and physiological parameters. No free text data are included. Further details can be found under information governance and ethics.

### Study Population

We included all patients who were: alive; aged 18-120, and registered with a TPP practice. Index of multiple deprivation,^15^ and rurality classification^16^ were derived for each patient based on their address. We excluded patients missing age, sex, index of multiple deprivation (IMD), and rurality classification, since their omission may indicate generally poor data quality.

We defined patients on each DMARD as those recorded with at least two prescriptions (one issued within 3 months before the search date, and the other between 3 to 6 months before the search date). Three-month search windows for prescriptions were chosen because in England GPs issue 93% of repeat prescriptions for a duration of three months or less.^17^

Azathioprine, leflunomide, methotrexate were chosen as a representative sample of DMARDs. These three medications are consistently classified as shared care across England, which avoids introducing potential confounding in the analysis of monitoring rates from inconsistent shared care statuses. We created codelists for each DMARD using NHS [dictionary of medicines and devices](https://www.nhsbsa.nhs.uk/pharmacies-gp-practices-and-appliance-contractors/dictionary-medicines-and-devices-dmd) (dm+d).

### Study Measures & Statistics

Monitoring tests relevant to the prescribed medication were selected (Table 1). These included full blood counts (FBC), liver function tests (LFT), urea and electrolytes (U&E), and blood pressure (BP). We excluded weight (for leflunomide) because it is not consistently listed as a monitoring requirement in all sources; namely the British National Formulary and Summary of Product Characteristics. Moreover, BP and weight checks would usually be carried out at the same time, so BP can be considered broadly representative of ‘physical’ health checks.

We selected patient groups which were relevant to health inequality priorities in England,^18^ or deemed susceptible to variation in monitoring. We grouped age into seven categories (18-29, 30-39, 40-49, 50-59, 60-69, 70-79, ≥80 years), sex into male or female, ethnicity into five categories (Black, Mixed, South Asian, White, Other), region into nine categories (East, East Midlands, London, North East, North West, South East, South West, West Midlands, Yorkshire and The Humber), rural-urban into eight categories (1: ‘most urban’, to 8: ‘most rural’),^16^ and IMD into five quintiles. Dementia, learning disability, serious mental illness, care home status, and housebound status were grouped as binary characteristics.

Codelists were specified to represent monitoring tests and patient groups using SNOMED CT. Each monitoring test typically represents a set of tests conducted together, therefore a single representative test was chosen (Table S1). For example, where Full Blood Counts are recommended we looked for the presence of a ‘Red blood cell count’ code.

Patients were defined as having missed monitoring if any of the relevant tests required for their given drug were not recorded in the last 3 months. Indicators (see Table S2) expressed the proportion of patients deemed to have missed monitoring (numerator), of all relevant patients prescribed the DMARD(s) being assessed (denominator). Higher percentages represent poorer monitoring performance. Each indicator was specified in analytic code within the OpenSAFELY framework, and was calculated from patient counts rounded to the nearest 5 to ensure anonymity.

#### Trend in DMARD monitoring rates

The numerator and denominator were generated for each indicator at monthly intervals between November 2019 and July 2022, then percentages calculated for each time-period. Note, each month in our results reflects a 3-month rolling average corresponding to data for the named month and the two months preceding it. For example, ‘May 2020’ represents patients not having tests 01/03/20 to 31/05/20. Time-periods of interest are defined in Table 2.

**Table 2.**
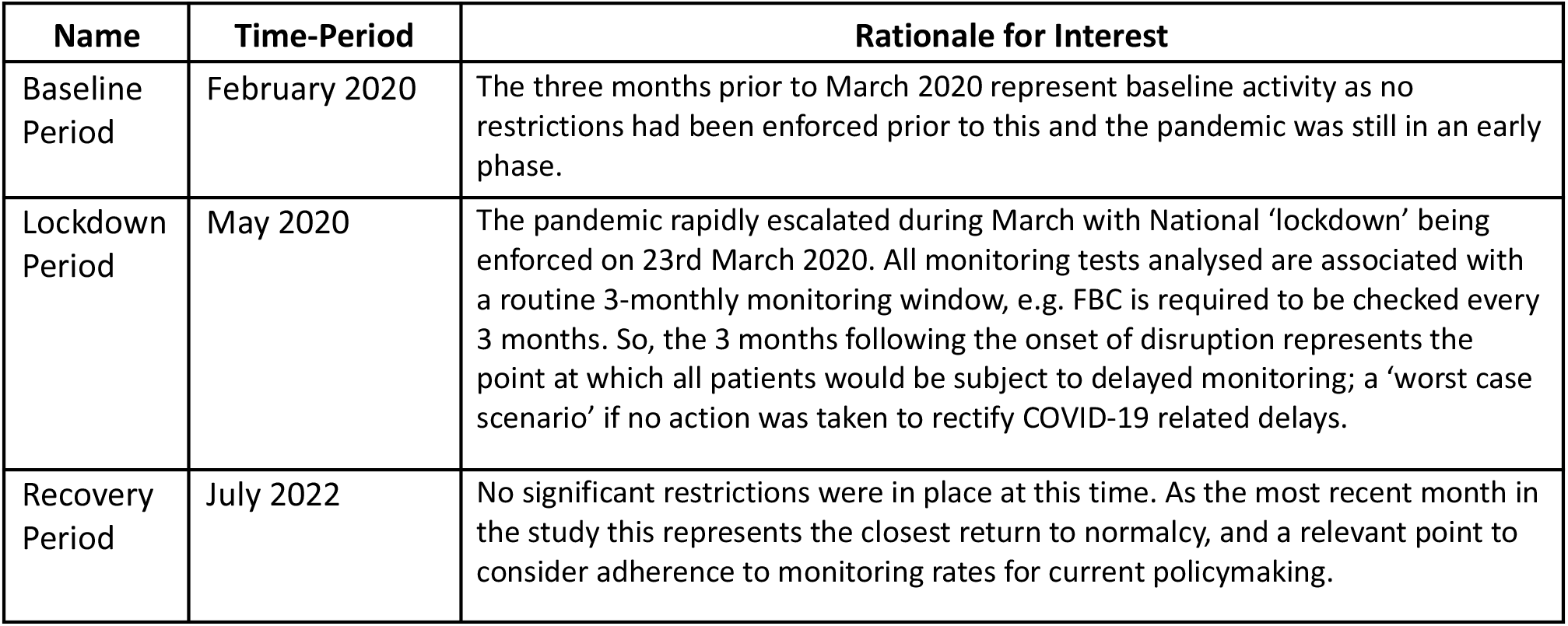
Time-periods of interest in relation to DMARD monitoring during the COVID-19 pandemic.

Total counts of the numerator and denominator were calculated for each indicator across the full study period. In this cumulative data, repeated events were counted for each time-period in which the event occurs, e.g. if a patient missed monitoring in two separate time-periods this was represented as two separate events. To report the extent of monitoring being repeatedly missed for the same patients, we also calculated the ratio of total missed monitoring events to unique patients with missed monitoring events across the full study period.

*t*-tests were conducted to investigate whether a significant change in monitoring rate occurred between February–May 2020.

#### Variability of change across GP practices

The indicator percentages for overall DMARD missed monitoring rate were summarised as deciles across all GP practices and presented as a decile chart.

#### Factors associated with change in monitoring rate

Cochran’s Q heterogeneity tests were conducted within each demographic/clinical subgroup to check for associations between patient factors and changes in monitoring rate.

### Software and Reproducibility

Data management and analysis was performed using the OpenSAFELY software libraries and Python, both implemented using Python 3·8. Inferential statistics were performed using R. This analysis was delivered using federated analysis, which involves carrying out patient-level analysis in multiple secure datasets, then later combining them: codelists and code for data management and analysis were specified once using the OpenSAFELY tools; then securely transmitted to the OpenSAFELY-TPP platform for execution against local patient data within TPP’s secure environment. Summary results were reviewed for disclosiveness and released for final outputs. All code for the OpenSAFELY platform for data management, analysis and secure code execution is shared for review and re-use under open licences at github.com/OpenSAFELY, or for this study at github.com/opensafely/Shared-Care-Monitoring.

### Patient and Public Involvement

Ensuring patient, professional and public trust is vital. Maintaining trust requires being transparent about how OpenSAFELY works, and ensuring patient voices are represented when designing research, analysing findings, and considering implications. Our website https://opensafely.org/ provides a detailed description of the platform in language suitable for a lay audience. We have participated in citizen juries exploring public trust in OpenSAFELY;^19^ we are currently co-developing an explainer video; we have ‘expert by experience’ patient representation on our OpenSAFELY Oversight Board; we have partnered with Understanding Patient Data to produce lay explainers on the importance of large datasets for research; we have presented at several public engagement events. We also work closely with appropriate medical research charities.

## Results

In the last month of the study (July 2022), 94,611 patients were identified as being regularly prescribed azathioprine, leflunomide or methotrexate. Of these patients, 60·9% were aged ≥60 years, 60·9% were female, and 91·1% were White. Table 3 shows their full demographic and clinical characteristics.

**Table 3.** Cohort description for patients who were included in at least one DMARD denominator at the end of the study period (July 2022).

| Characteristic | Category | n | % * |
| --- | --- | --- | --- |
| Population | Total | 94,611 | 100.0% |
| Age Band | 18-29 | 3,465 | 3.7% |
|  | 30-39 | 5,905 | 6.2% |
|  | 40-49 | 9,555 | 10.1% |
|  | 50-59 | 18,050 | 19.1% |
|  | 60-69 | 22,410 | 23.7% |
|  | 70-79 | 23,490 | 24.8% |
|  | 80+ | 11,730 | 12.4% |
| Sex | Female | 57,620 | 60.9% |
|  | Male | 36,990 | 39.1% |
| Ethnicity | Black | 1,050 | 1.1% |
|  | Mixed | 625 | 0.7% |
|  | Other | 750 | 0.8% |
|  | South Asian | 5,180 | 5.5% |
|  | White | 86,215 | 91.1% |
|  | Missing | 790 | 0.8% |
| Region | East | 23,105 | 24.4% |
|  | East Midlands | 16,365 | 17.3% |
|  | London | 3,040 | 3.2% |
|  | North East | 5,040 | 5.3% |
|  | North West | 7,860 | 8.3% |
|  | South East | 6,605 | 7.0% |
|  | South West | 15,260 | 16.1% |
|  | West Midlands | 2,925 | 3.1% |
|  | Yorkshire and The Humber | 14,140 | 15.0% |
|  | Missing | 270 | 0.3% |
| Care Home | Yes | 575 | 0.6% |
|  | No | 94,035 | 99.4% |
| Dementia | Yes | 1,370 | 1.5% |
|  | No | 93,245 | 98.6% |
| Housebound | Yes | 3,380 | 3.6% |
|  | No | 91,235 | 96.4% |
| Learning Disability | Yes | 375 | 0.4% |
|  | No | 94,235 | 99.6% |
| Serious Mental Illness | Yes | 940 | 1.0% |
|  | No | 93,670 | 99.0% |
| Index of Multiple Deprivation | 1st quintile (most deprived) | 15,160 | 16.0% |
|  | 2nd quintile | 17,555 | 18.6% |
|  | 3rd quintile | 21,210 | 22.4% |
|  | 4th quintile | 21,075 | 22.3% |
|  | 5th quintile (least deprived) | 19,610 | 20.7% |
| Rural-Urban Classification | Urban major conurbation | 14,435 | 15.3% |
|  | Urban minor conurbation | 6,155 | 6.5% |
|  | Urban city and town | 49,290 | 52.1% |
|  | Urban city and town in a sparse setting | 235 | 0.3% |
|  | Rural town and fringe | 13,180 | 13.9% |
|  | Rural town and fringe in a sparse setting | 630 | 0.7% |
|  | Rural village and dispersed | 9,855 | 10.4% |
|  | Rural village and dispersed in a sparse setting | 835 | 0.9% |

\*patient counts for subgroups were rounded to the nearest 5, so percentages were calculated as a proportion of the subgroup total.

### Trend in DMARD monitoring rates

Throughout the study, patients missed monitoring at an average rate of 31·1% (977,354 of 3,146,849 prescribing events). The ratio of missed monitoring events to unique patients was 8·3. Between the baseline and lockdown periods, there was a significant rise in missed monitoring rates across the population, increasing from 28·4% to 40·8% (+12·4 percentage points, *p* <0·001). However, between the lockdown and recovery periods, rates decreased to 28·1% (-12·7 percentage points). For a detailed breakdown of results by DMARD and monitoring test, see Table S3. For breakdowns by demographic and clinical subgroups (including statistical analysis of change over time), see Table S4.

#### Medication Type

The percentage of patients identified as missing monitoring was greatest for leflunomide (65·0%; 125,850 of 193,665 patients, ratio 13·5 events per unique patient) and lowest for methotrexate (24·6%; 517,770 of 2,106,050 patients, ratio 7·0 events per unique patient).

All DMARDs exhibited an increase in missed monitoring rates immediately following the onset of lockdown (April–June 2020; Figure 1). These rates showed considerable recovery through July–August 2020. The increase in missed monitoring rates between the baseline and lockdown period was similar for methotrexate (+11·7 percentage points) and azathioprine (+12·3 percentage points), whereas a noticeably greater change was seen for leflunomide (+20·7 percentage points). By the recovery period, monitoring rates had returned to close to baseline period, in the case of azathioprine (38·5% baseline and 35·5% recovery), methotrexate (21·7% baseline and 22·3% recovery), leflunomide (56·1% baseline and 57·9% recovery).

**Figure 1.**
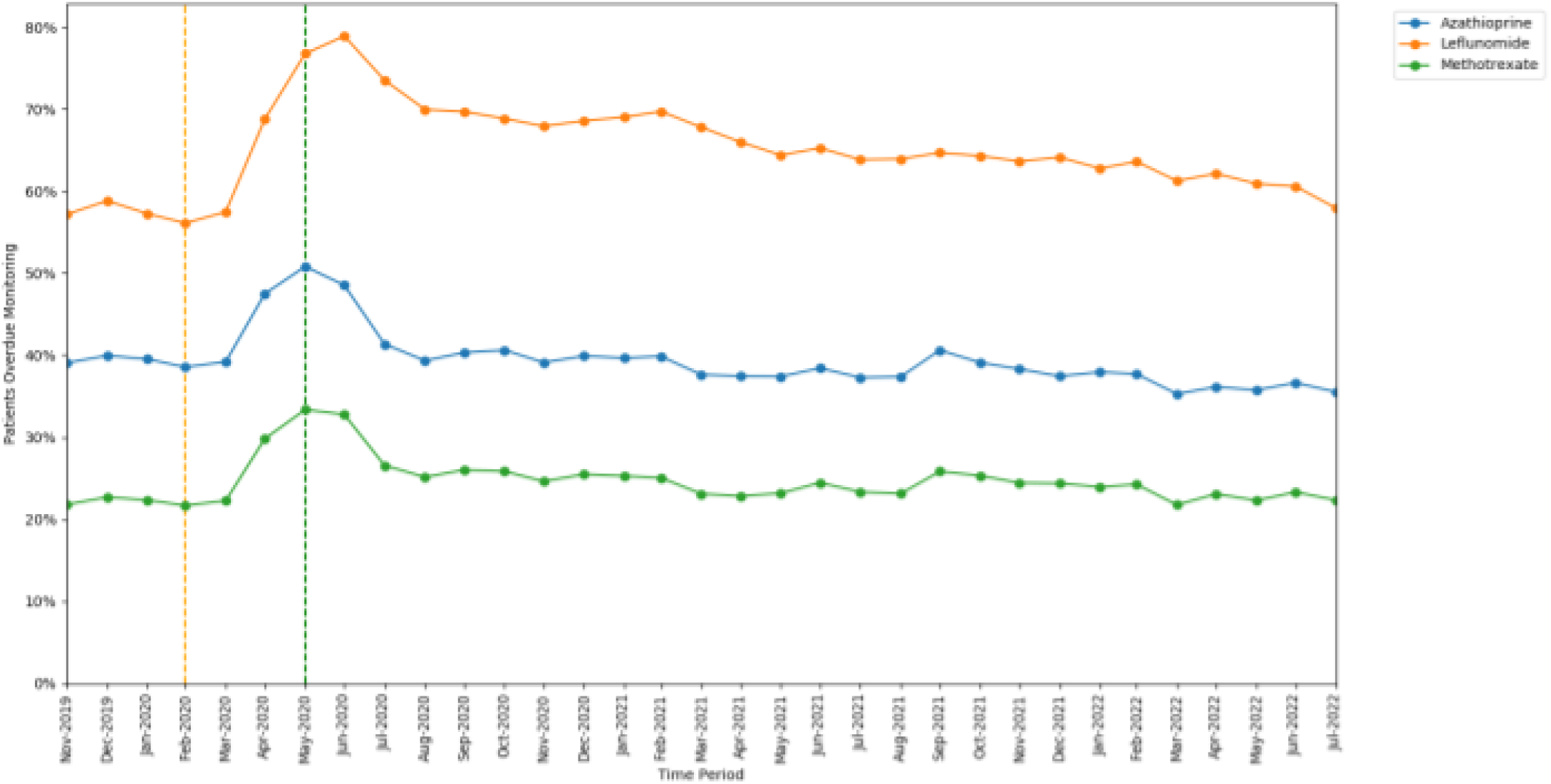
Proportion of patients overdue monitoring between November 2019 to July 2022, broken down by shared care medication. The baseline period before lockdown is shown as an orange dashed vertical line. The monitoring window, measured as 3 months from the onset of the March 2020 COVID-19 lockdown, is shown as a green dashed vertical line.

#### Monitoring Type

The monitoring test with the highest rate of missed monitoring was BP (57·4%; 111,215 of 193,665 patients, ratio 12·3 events per unique patient). All other tests had very similar rates: LFT (25·2%; 793,870 of 3,146,849 patients, ratio 7·0 events per unique patient), FBC (26·1%; 820,895 of 3,146,849 patients, ratio 7·3 events per unique patient), U&E (25·7%; 810,255 of 3,146,849 patients, ratio 7·2 events per unique patient).

All monitoring tests exhibited an increase in missed monitoring immediately following the onset of lockdown (April–June 2020; Figure 2). The increase in missed monitoring rates between the baseline and lockdown period was similar for FBC (+12·61 percentage points), LFT (+12·75 percentage points), and U&E (+12·66 percentage points), whereas a greater change was seen for BP (+24·46 percentage points). All rates showed considerable recovery through July–August 2020. By the recovery period most tests had returned to similar rates as at the baseline period: FBC (23·17% baseline and 23·58% recovery), LFT (22·04% baseline and 23·05% recovery), U&E (22·83% baseline and 23·59% recovery), with BP remaining slightly higher than baseline (45·75% baseline and 49·88% recovery).

**Figure 2.**
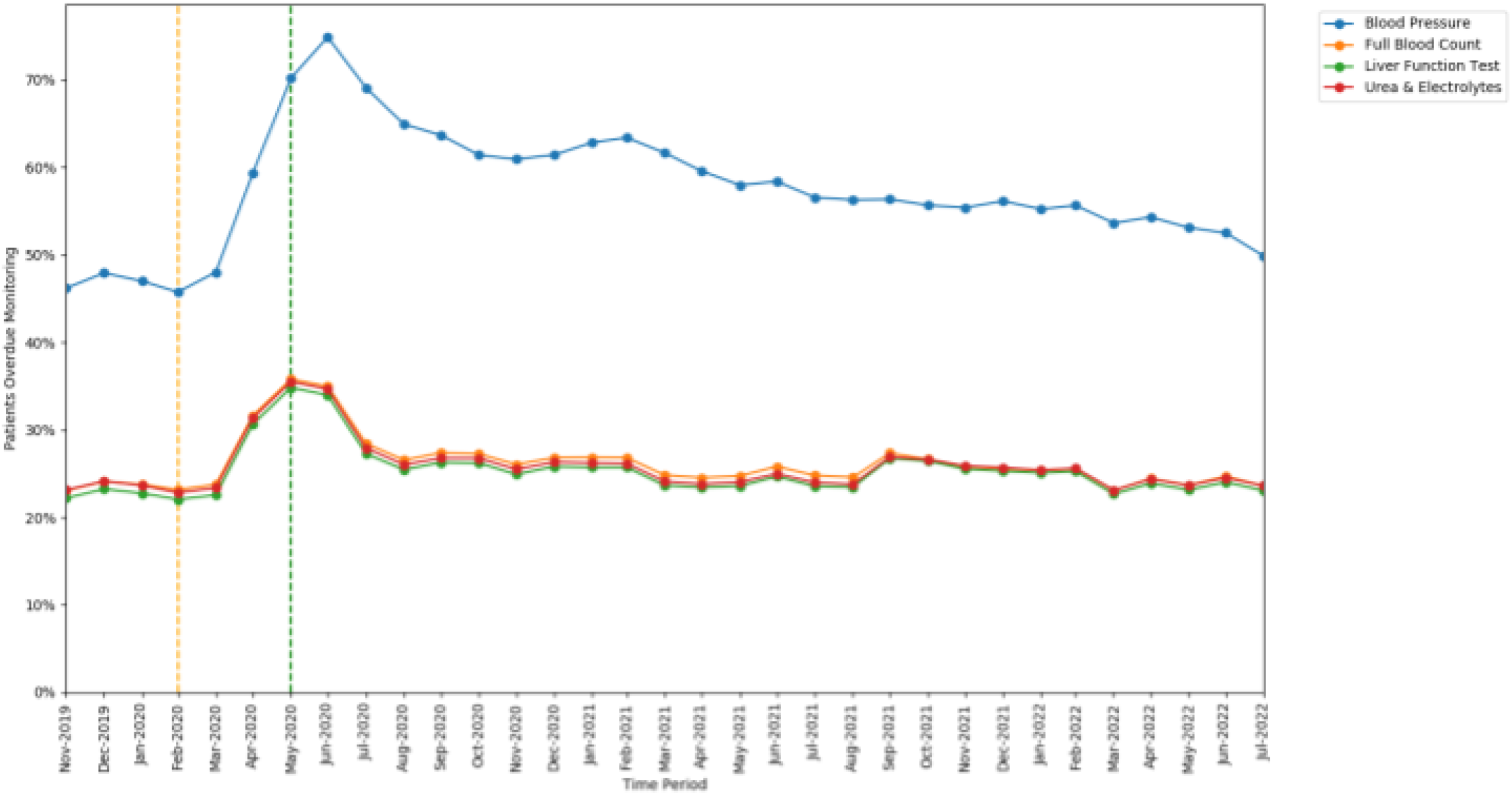
Proportions of patients overdue DMARD monitoring between November 2019 to July 2022, broken down by monitoring test type. The baseline period before lockdown is shown as an orange dashed vertical line. The monitoring window, measured as 3 months from the onset of the March 2020 COVID-19 lockdown, is shown as a green dashed vertical line.

### Variability of change across GP practices

At the baseline period (February 2020), the median missed monitoring rate across GP practices was 25.0%. There was a large interdecile range, from 8·3% in the 1st decile to 63·3% in the 9th decile (Figure 3).

**Figure 3.**
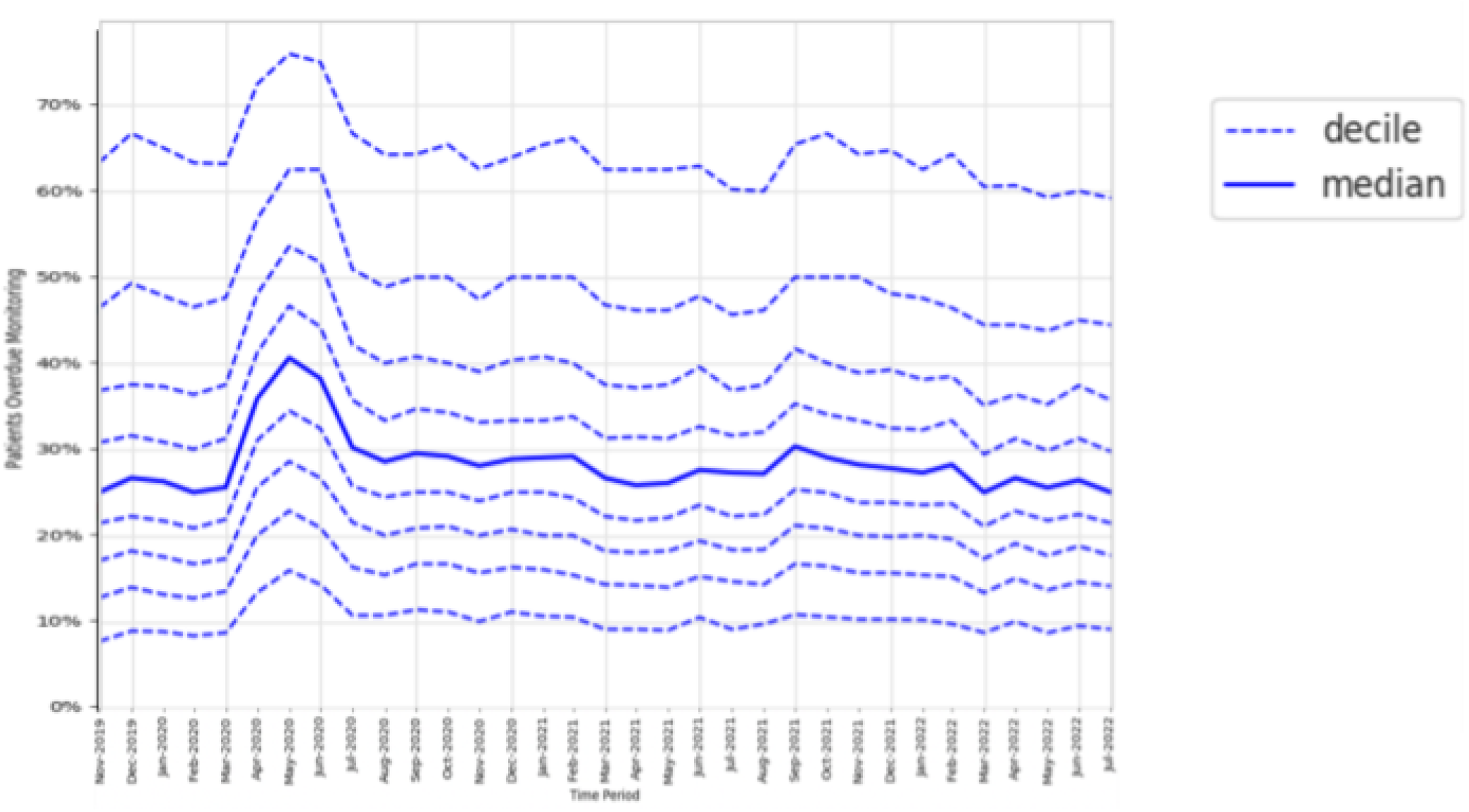
Practice level decile charts for proportions of patients overdue DMARD monitoring between November 2019 to July 2022.

At the lockdown period (May 2020), the median missed monitoring rate was 40·6% (+15·6 percentage points from baseline). The interdecile range widened slightly with the 9th decile increasing by +12·6 percentage points from baseline, compared to +7·6 percentage points for the 1st decile.

At the recovery period (July 2022), the interdecile range had narrowed compared to baseline.

### Factors associated with change in monitoring rate

Mean monitoring rates at baseline, lockdown, and recovery periods are presented for each group of patient characteristics in Table S4.

All groups showed significant increase in missed monitoring rates between the baseline and lockdown period. This increase differed for: age-group categories (heterogeneity test: Cochran’s Q=37·526, *p*<0·001; Figure 4), ranging from +8·3 percentage points in the 18-29 years age-group to +13·7 percentage points in the 70-79 years age-group; sex categories, (Q=4·5, *p*=0·034; Figure 5), ranging from +11·9 percentage points in males to +12·8 percentage points in females; and regions (Q=78·869, *p*<0·001; Figure 6), ranging from +9·2 percentage points in the North East to +17·0 percentage points in the North West.

**Figure 4.**
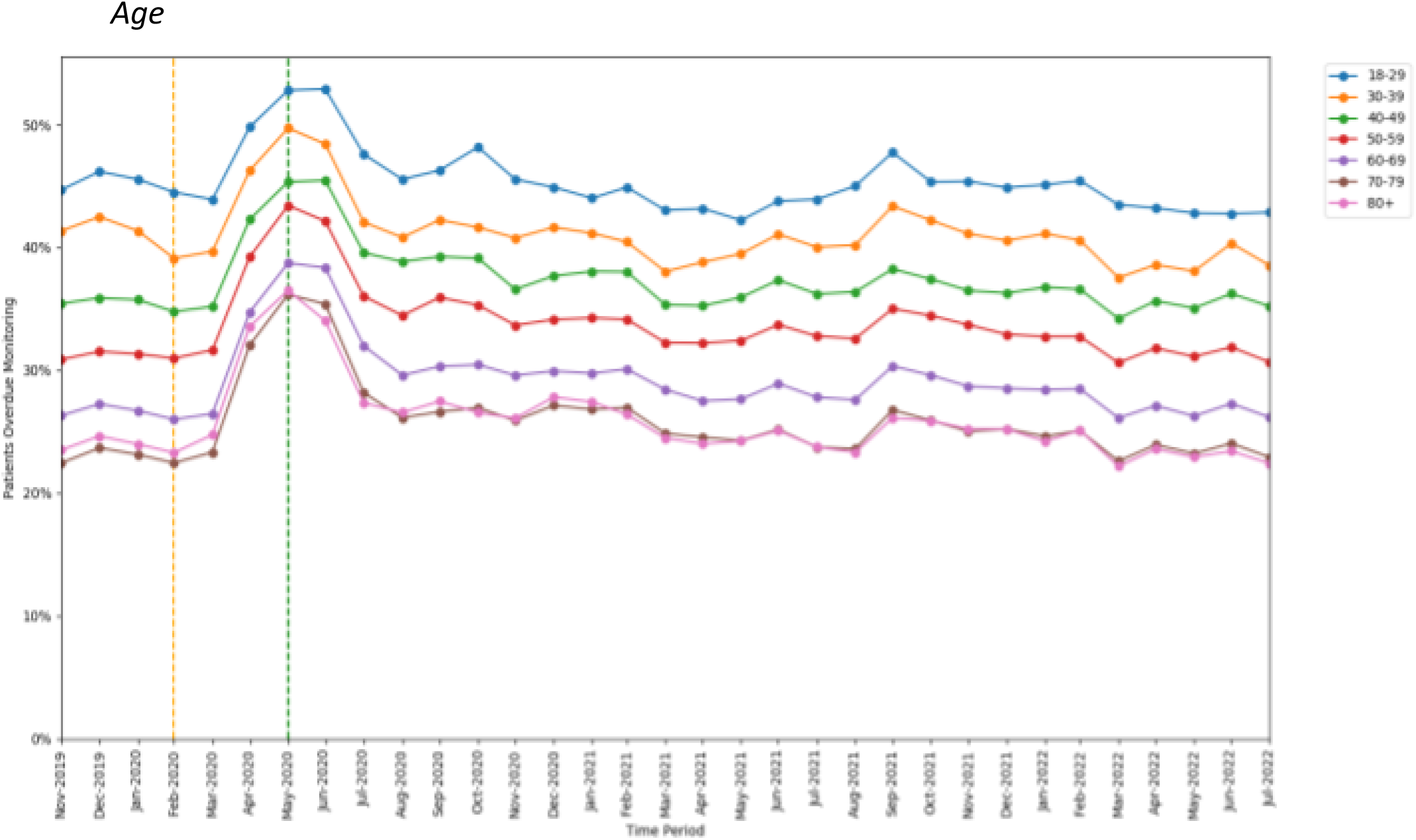
Proportions of patients overdue DMARD monitoring between November 2019 to July 2022, broken down by age-band. The baseline period before lockdown is shown as an orange dashed vertical line. The monitoring window, measured as 3 months from the onset of the March 2020 COVID-19 lockdown, is shown as a green dashed vertical line.

**Figure 5.**
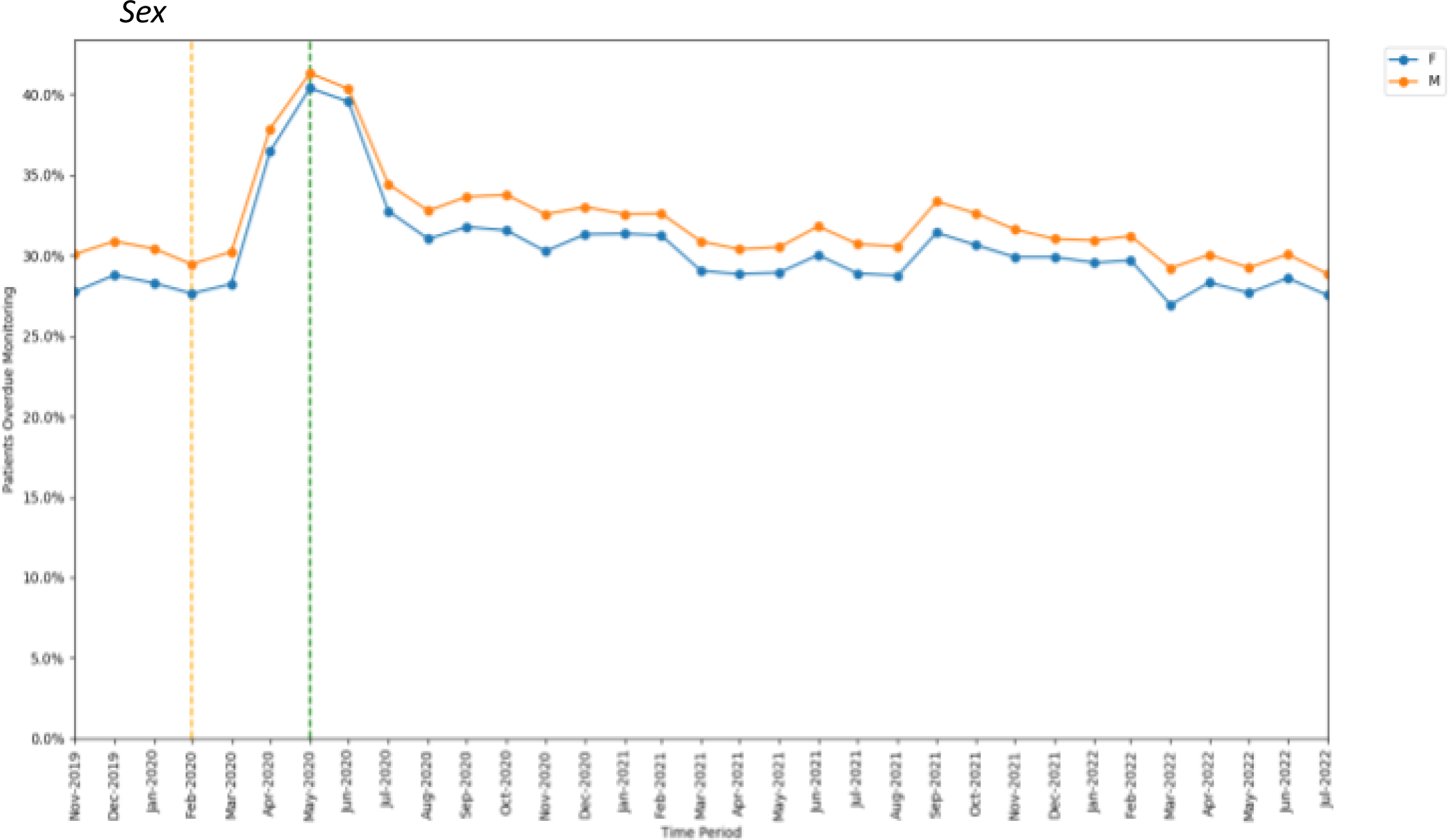
Proportions of patients overdue DMARD monitoring between November 2019 to July 2022, broken down by sex. The baseline period before lockdown is shown as an orange dashed vertical line. The monitoring window, measured as 3 months from the onset of the March 2020 COVID-19 lockdown, is shown as a green dashed vertical line.

**Figure 6.**
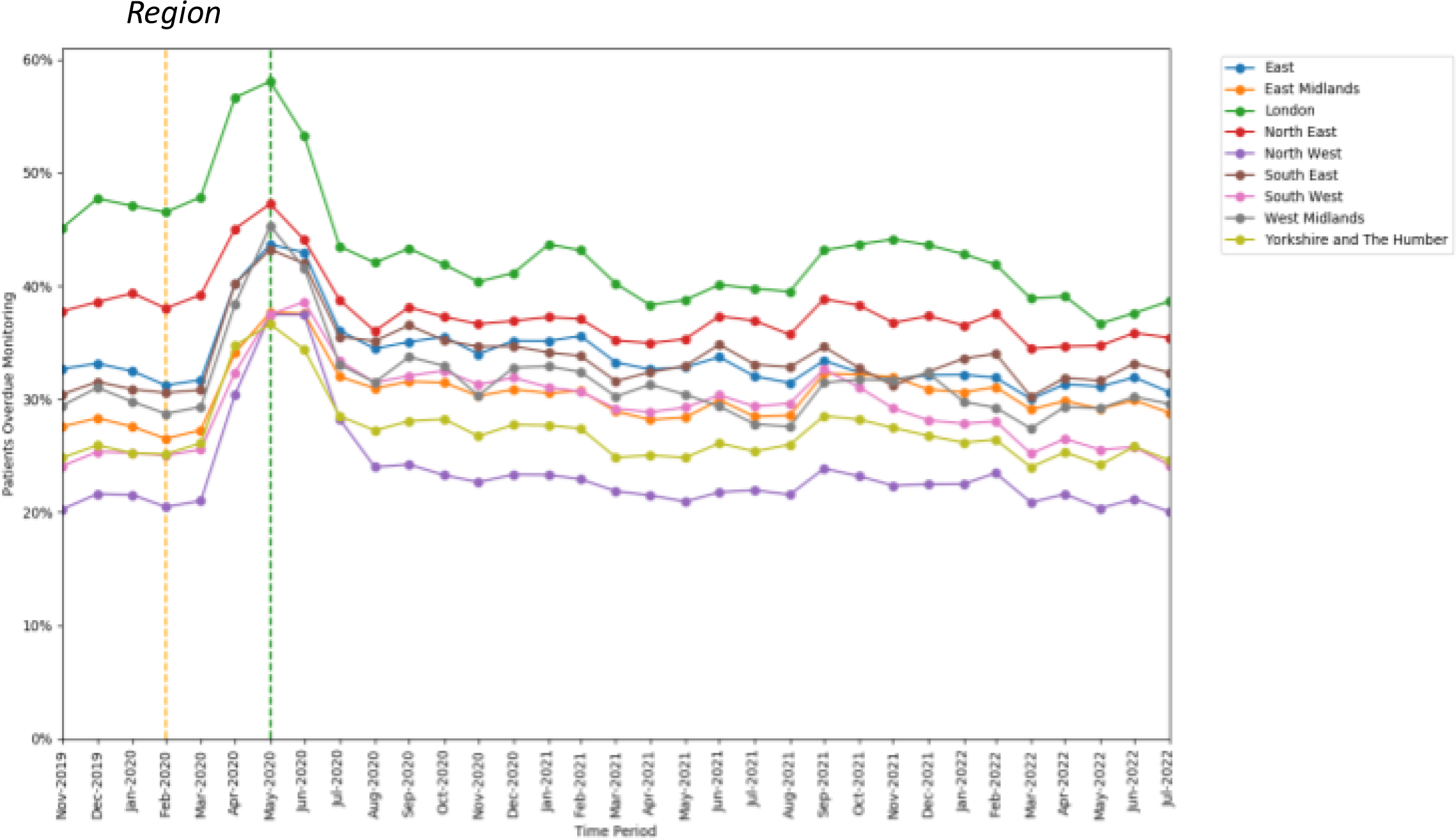
Proportions of patients overdue DMARD monitoring between November 2019 to July 2012, broken down by region. The baseline period before lockdown is shown as an orange dashed vertical line. The monitoring window, measured as 3 months from the onset of the Mareh 2020 COVID-19 Iockdown, is shown as a green dashed vertical line.

Throughout the study, substantial overall differences were apparent in missed monitoring rates within the groups of age, sex, region (Figures 4 to 6), and also ethnicity, learning disabilities, serious mental illness, deprivation, and rural-urban score (Figures S1 to S5).

## Discussion

### Summary

A significant increase in missed safety monitoring was observed following COVID-19 lockdown measures in March 2020, suggesting that lockdowns negatively affect the ability for patients to undergo monitoring. Deterioration in monitoring was disproportionately large for: BP testing (a unique requirement for leflunomide), older people, females, and specific regions. Safety monitoring recovered rapidly across all groups as lockdown measures were eased.

Throughout the study period, DMARD safety monitoring fell below the recommended standard, with patients missing monitoring at an average rate of 31·1%, especially in certain groups such as younger people, ethnic minorities, specific regions, patients living in more deprived or urban areas, and patients with an SMI or learning disability. This raises questions relating to specific health inequalities, and more generally whether current monitoring requirements are appropriate or sustainable for all patients.

### Strengths and Limitations

The scale and comprehensiveness of data in the OpenSAFELY platform is greater than any other method for accessing GP data. Historically, attempts to audit monitoring compliance in primary care would have typically relied on manual audit within a practice. By contrast, OpenSAFELY allows efficient analyses using data which is broadly representative of the English population, providing a national overview of monitoring compliance.^20^ A second strength is the study’s reproducibility and transparency. The complete set of code for the platform and all data curation/analysis from raw data to final output, is shared publicly on GitHub for peer review and efficient reuse.

We note some limitations. Firstly, our data only includes results from tests carried out in primary care, or carried out in secondary care where results are returned to GPs as structured data. Test results performed outside GP practices are omitted from our data unless they are communicated to the GP practice and coded as structured data by GP practice staff. In most circumstances, where GP practices prescribe the medications they will usually also perform the monitoring. Secondly, in some instances a clinician or the patient may have decided to stop the medication since the prescription issue date (especially during the pandemic months), which may invalidate the requirement for monitoring to be completed in the expected time-period. Both of these limitations may lead to some patients falsely appearing overdue for monitoring. Conversely, some tests may have been counted that were carried out for reasons unrelated to DMARD monitoring. We were also unable to capture prescriptions issued by secondary care, this data has historically been challenging to obtain.^21^ Recently, NHS Digital have made prescription data available for some hospitals - we will aim to incorporate this in future research.^22^ Lastly, we report data for three DMARDs, therefore findings may not be generalisable for other high-risk drugs.

### Comparison with Existing Literature

This study is the first to comprehensively evaluate DMARD monitoring at a national scale during the pandemic. Our findings are consistent with previous OpenSAFELY research which identified deterioration in monitoring across a range of other medications and monitoring tests following the March 2020 lockdown.^11, 12^ For example, Fisher *et al.* (2023)^12^ reported 19·6% of patients on methotrexate missed LFT on average between January to March 2020, whilst in the current study we reported 22·1% of patients on methotrexate missed LFT, FBC, or U&E across the same time-period. This small difference is to be expected as our Fisher *et al.* study used indicators specific to single tests, whilst the current study included all relevant tests in a combined indicator. The difference may also be partially attributable to our study searching for a narrower range of key monitoring test codes, giving a stricter view of what is acceptable monitoring.

Research on the determinants of monitoring adherence is sparse, but parallels can be drawn with research on the determinants of medication-taking adherence. The differences we identified in monitoring rates between certain patient groups are consistent with research suggesting that patient-related factors (notably age, sex, ethnicity, cognitive function, and socio-economic status) can influence medication-taking adherence.^23^ Although, it should be noted that many patient-related factors have been reported as having an inconsistent impact, suggesting that determinants of adherence are multifaceted and context-specific.^24, 25^

### Policy Implications and Interpretation

This research highlights that DMARD monitoring is often not adhered to. However, wide variance in monitoring rates between practices indicates that some practices have been able to implement successful strategies. This presents opportunities for sharing best practice, which NHS commissioners may be able to facilitate at scale.

The deterioration that this study observed in DMARD monitoring during lockdown, and that other studies have observed in routine services,^11^ offers insight for health policymakers regarding the effects of the pandemic and its management strategies. A combination of factors likely contributed to this deterioration including: diversion of primary care resources towards managing COVID-19, guidance from national bodies to enable reduced monitoring frequency, supply problems with blood sample bottles, and patients choosing to avoid healthcare for fear of exposure to COVID-19. This understanding will help inform assessment of the risks and benefits of lockdown measures in future pandemics, and how the impact on groups which were more severely affected could be mitigated.

The disproportionately large deterioration observed in BP monitoring may suggest that clinicians deemed it less critical than blood tests, reflecting appropriate prioritisation of monitoring. Regional differences in deterioration may suggest that regions prioritised DMARD monitoring differently during the pandemic. Factors underpinning regional differences may include the availability of local workforce and proximity of facilities to support the pandemic response; both have been suggested as relevant to service resilience during the pandemic.^26^ Differences in resilience may also be related to known variability in the robustness of local shared care processes and documentation,^27^ and regional differences which have been observed in the time between first rheumatoid arthritis review to DMARD initiation.^28^

The overall differences in monitoring rates observed between various groups highlight ongoing inequalities. For example, younger age-groups generally missed monitoring more often than older age-groups. This may indicate that younger age-groups exhibit decreased engagement with monitoring, which is consistent with previous research.^29^ Less monitoring in younger-age groups means there are fewer opportunities for corrective dose adjustments, which may help explain findings from a recent study; that younger patients taking methotrexate or azathioprine were more likely to have abnormal monitoring test results when tested.^30^ Although, it should be noted that older age-groups are more vulnerable to DMARD toxicity.^31^

Health commissioners should consider targeted strategies to reduce inequalities as appropriate, for example, addressing the disproportionately high rate of missed BP monitoring (often recurring in the same patients on leflunomide) may be a pragmatic starting point. Since the pandemic, standardised national processes for shared care have been published by NHSE in relation to several medicines,^32^ adopting these national standards should help local health commissioners reduce inequalities.

This analysis has substantial implications for NHS data infrastructure. Historically, practical and privacy challenges around accessing primary care data meant that national audits relied upon manual data collection by local teams and central collation - an approach with high resource and time costs. The OpenSAFELY platform enabled us to execute a single analysis for a representative sample of the population in near-real-time (with data available in OpenSAFELY only 2-9 days after being entered by a clinician) whilst leaving data in situ. This efficient approach allows our analysis to be easily extended (by building upon open-access code) to wider populations, or to provide more granular data on demographic or clinical subgroups. Ultimately, OpenSAFELY provides tools which facilitate rapid detailed feedback to NHS commissioners and clinicians, enabling timely and meaningful interventions to improve patient outcomes.

### Future Research

To inform policy interventions, further research is needed to explore the causes of poorer performance amongst specific practices and patient groups. This could be done through qualitative studies surveying/interviewing those with poorer monitoring rates to identify the individual traits, attitudes, behaviours, and circumstances underpinning their poorer monitoring adherence.

More broadly, the pandemic offers an unprecedented opportunity of a natural experiment, since changes to clinical practice (such as the frequency of safety monitoring) have occurred that would have been unethical in normal circumstances. This allows researchers to test whether certain principles of clinical practice could be optimised. Recent research has suggested that the clinical impact of missing DMARD monitoring is minimal, with one study suggesting that reducing the frequency of methotrexate monitoring from 3 to 6 monthly did not increase abnormal test results nor cause harm to patients.^33^ However, such analysis is easily confounded by patient-specific factors related to clinical risk, which influence the likelihood of monitoring occurring. For example, patients at lower risk of negative outcomes may be less motivated to attend monitoring, or less proactively followed up by clinicians. By contrast, patients at higher risk may be more likely to have accessibility issues inhibiting monitoring, or more reluctant to attend monitoring due to fear of contracting COVID-19 with their clinical vulnerability. Consequently, patients who miss monitoring may have an intrinsically different likelihood of negative outcomes. Studies could attempt to evaluate the clinical impact of reduced monitoring frequency within patient groups stratified by clinical risk factors, and break results down for various medications/tests. This may reveal whether certain medications or patient groups benefit from monitoring to different extents, informing more tailored cost-effective strategies for who should receive which monitoring tests, and how often.

### Conclusion

A transient deterioration in DMARD safety monitoring occurred across all groups during the COVID-19 pandemic, acutely around the time of lockdown measures with performance mostly recovering rapidly as lockdown measures were eased. Long-term differences in safety monitoring rates exist between several sociodemographic and clinical subgroups, and the pandemic impacted monitoring in these populations to varying extents. Health commissioners should identify causes of poorer monitoring in specific groups and consider implementing supportive measures. Monitoring rates varied substantially between practices, suggesting opportunities exist to improve service consistency by sharing good practice and adopting standardised shared care processes. These analyses demonstrate the capability of the OpenSAFELY platform as an effective tool to provide actionable insights on health service provision and inequalities.

## Data Availability

All data were linked, stored and analysed securely within the OpenSAFELY platform https://opensafely.org/. All code is shared openly for review and re-use under MIT open license. Detailed pseudonymised patient data is potentially re-identifiable and therefore not shared. We rapidly delivered the OpenSAFELY data analysis platform without prior funding to deliver timely analyses on urgent research questions in the context of the global Covid-19 health emergency: now that the platform is established we are developing a formal process for external users to request access in collaboration with NHS England; details of this process are available at [OpenSAFELY.org](http://opensafely.org/).
All clinical and medicines codelists are openly available for inspection and reuse at https://codelists.opensafely.org/.

https://github.com/opensafely/Shared-Care-Monitoring

## Administrative

## Acknowledgements

We are very grateful for all the support received from the TPP Technical Operations team throughout this work, and for generous assistance from the information governance and database teams at NHS England and the NHS England Transformation Directorate.

## Information governance and ethical approval

NHS England is the data controller; TPP is the data processor; and the researchers on OpenSAFELY are acting with the approval of NHS England. This implementation of OpenSAFELY is hosted within the TPP environment which is accredited to the ISO 27001 information security standard and is NHS IG Toolkit compliant;^34,35^ patient data has been pseudonymised for analysis and linkage using industry standard cryptographic hashing techniques; all pseudonymised datasets transmitted for linkage onto OpenSAFELY are encrypted; access to the platform is via a virtual private network (VPN) connection, restricted to a small group of researchers; the researchers hold contracts with NHS England and only access the platform to initiate database queries and statistical models; all database activity is logged; only aggregate statistical outputs leave the platform environment following best practice for anonymisation of results such as statistical disclosure control for low cell counts.^36^ The OpenSAFELY research platform adheres to the obligations of the UK General Data Protection Regulation (GDPR) and the Data Protection Act 2018. In March 2020, the Secretary of State for Health and Social Care used powers under the UK Health Service (Control of Patient Information) Regulations 2002 (COPI) to require organisations to process confidential patient information for the purposes of protecting public health, providing healthcare services to the public and monitoring and managing the COVID-19 outbreak and incidents of exposure; this sets aside the requirement for patient consent.^37^ Taken together, these provide the legal bases to link patient datasets on the OpenSAFELY platform. GP practices, from which the primary care data are obtained, are required to share relevant health information to support the public health response to the pandemic, and have been informed of the OpenSAFELY analytics platform.

This study was approved by the Health Research Authority (REC reference 20/LO/0651) and by the LSHTM Ethics Board (reference 21863).

## Data sharing

Access to the underlying identifiable and potentially re-identifiable pseudonymised electronic health record data is tightly governed by various legislative and regulatory frameworks, and restricted by best practice. The data in OpenSAFELY is drawn from General Practice data across England where TPP is the Data Processor. TPP developers initiate an automated process to create pseudonymised records in the core OpenSAFELY database, which are copies of key structured data tables in the identifiable records. These pseudonymised records are linked onto key external data resources that have also been pseudonymised via SHA-512 one-way hashing of NHS numbers using a shared salt. Bennett Institute for Applied Data Science developers and PIs holding contracts with NHS England have access to the OpenSAFELY pseudonymised data tables as needed to develop the OpenSAFELY tools. These tools in turn enable researchers with OpenSAFELY Data Access Agreements to write and execute code for data management and data analysis without direct access to the underlying raw pseudonymised patient data, and to review the outputs of this code. All code for the full data management pipeline—from raw data to completed results for this analysis—and for the OpenSAFELY platform as a whole is available for review at github.com/OpenSAFELY.

## Guarantor

BG is the guarantor.

## Contributorship

Conceptualization: AB, REC, BMK

Data curation: AB, LF, LEMH, CC, SCJB, AM, GH, CB, JC, JP, FH, SH

Formal analysis: AB, LF, MW, WJH, HJC

Funding acquisition: BG

Investigation: AB, LF, MW, WJH, HJC

Methodology: AB, WJH, BMK

Project administration: AM

Resources: AM, SCJB, GH, CB, JC, JP, FH, SH

Software: SCJB, GH, CB, JC, JP, FH, SH

Supervision: ZK, AM, BG, BMK

Visualisation: AB, LF, BMK

Writing - original draft: AB

Writing - review & editing: AB, LF, HJC, MW, WJH, LEMH, CC, VS, REC, JBG, MDR, KB, ZK, RC, CW, AJW, ALS, SCJB, AM, GH, BG, BMK

## Supplementary Tables

**Table S1.** Key features which were assessed to determine the completion of monitoring tests, and links to codelists which identified these features.

| Monitoring Test | Representative Feature | Codelist |
| --- | --- | --- |
| Urea and electrolytes (U&E) | Sodium level | https://www.opencodelists.org/codelist/opensafely/sodium-tests/254b9f34/ |
| Full blood count (FBC) | Red blood cell count | https://www.opencodelists.org/codelist/opensafely/red-blood-cell-rbc-tests/576a859e/ |
| Liver function test (LFT) | Alanine-aminotransferase level | https://www.opencodelists.org/codelist/opensafely/alanine-aminotransferase-alt-tests/2298df3e/ |
| Blood pressure (BP) | Systolic blood pressure | https://www.opencodelists.org/codelist/opensafely/systolic-blood-pressure-qof/3572b5fb/ |

**Table S2.**
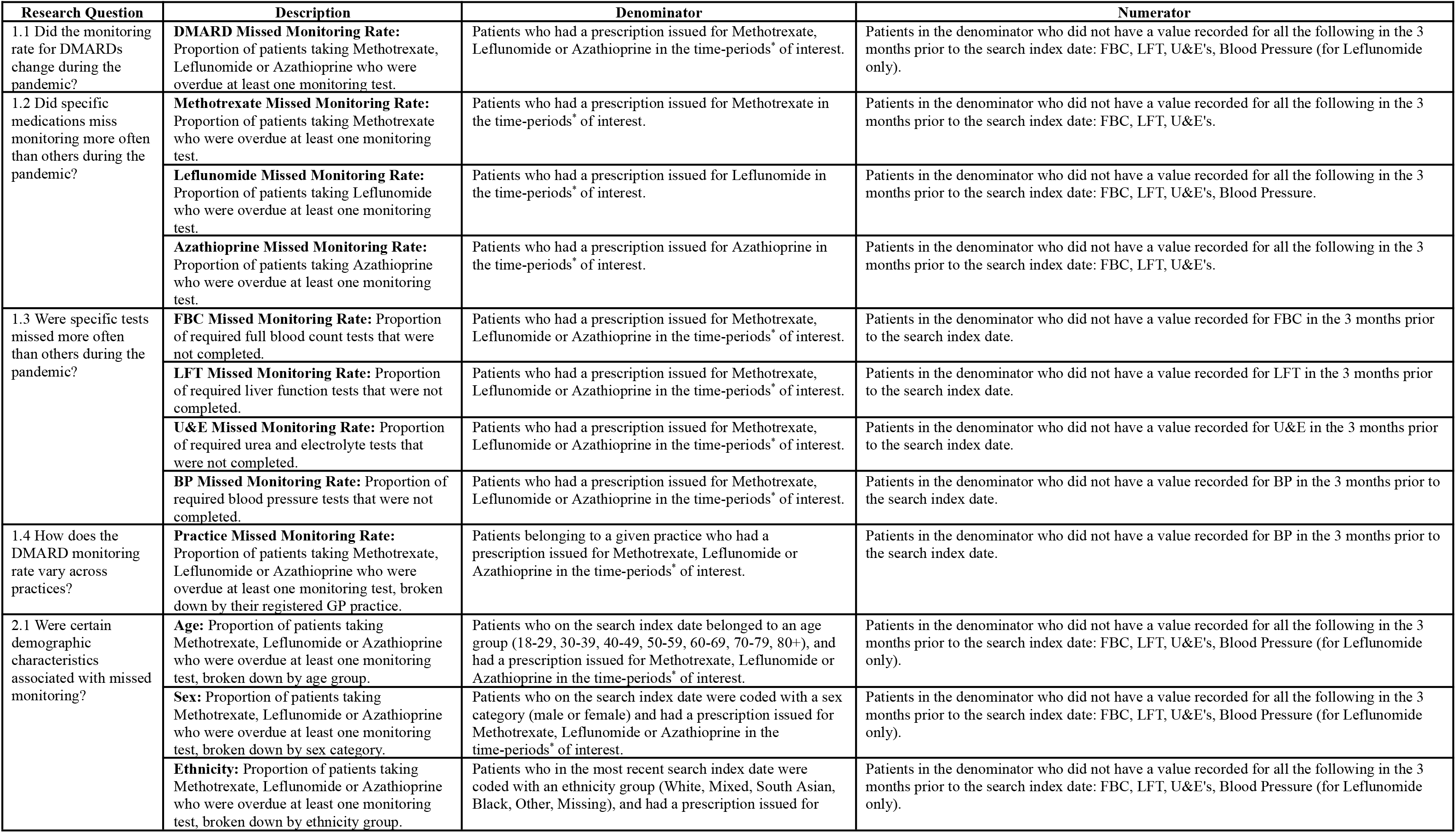
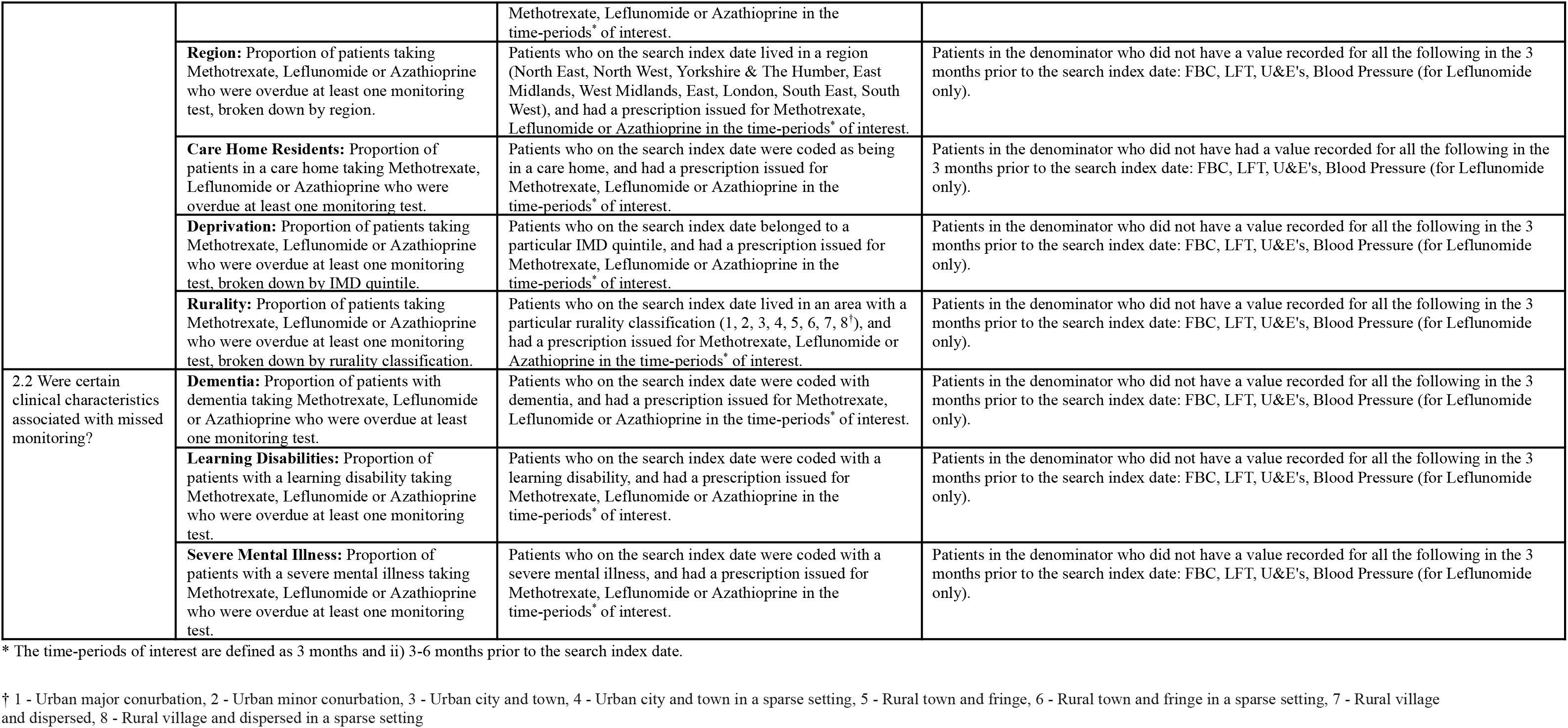
Indicator definitions.

**Table S3.** DMARD missed monitoring rates at baseline (Dec19 to Feb20), lockdown (Mar20 to May20) and recovery (May22 to Jul22), with cumulative counts of missed monitoring events and unique patients experiencing a missed monitoring event over the full study period (Nov19 to Jul22), broken down by medication and monitoring test.

| Category | Missed Monitoring Rate |  |  |  | Cumulative Count |  |  |  |
| --- | --- | --- | --- | --- | --- | --- | --- | --- |
|  | Baseline Period | Lockdown Period | Recovery Period | Impact (Lockdown – Baseline) | Denominator Events | Number of missed monitoring events (% of denominator) | Number of unique patients with a missed monitoring event | Ratio of missed monitoring events to unique patients with a missed monitoring event |
| Population | 28.4% | 40.8% | 28.1% | 12.4 | 3,146,849 | 977,354 (31.1%) | 117,784 | 8.3 |
| Methotrexate | 21.7% | 33.3% | 22.3% | 11.7 | 2,106,050 | 517,770 (24.6%) | 74,048 | 7.0 |
| Azathioprine | 38.5% | 50.8% | 35.5% | 12.3 | 847,125 | 333,725 (39.4%) | 34,892 | 9.6 |
| Leflunomide | 56.1% | 76.7% | 57.9% | 20.7 | 193,665 | 125,850 (65.0%) | 9,302 | 13.5 |
| FBC | 23.2% | 35.8% | 23.6% | 12.6 | 3,146,849 | 820,895 (26.1%) | 112,599 | 7.3 |
| LFT | 22.0% | 34.8% | 23.0% | 12.8 | 3,146,849 | 793,870 (25.2%) | 112,858 | 7.0 |
| U&E | 22.8% | 35.5% | 23.6% | 12.7 | 3,146,849 | 810,255 (25.7%) | 112,805 | 7.2 |
| BP | 45.7% | 70.2% | 49.9% | 24.5 | 193,665 | 111,215 (57.4%) | 9,008 | 12.3 |

**Table S4.**
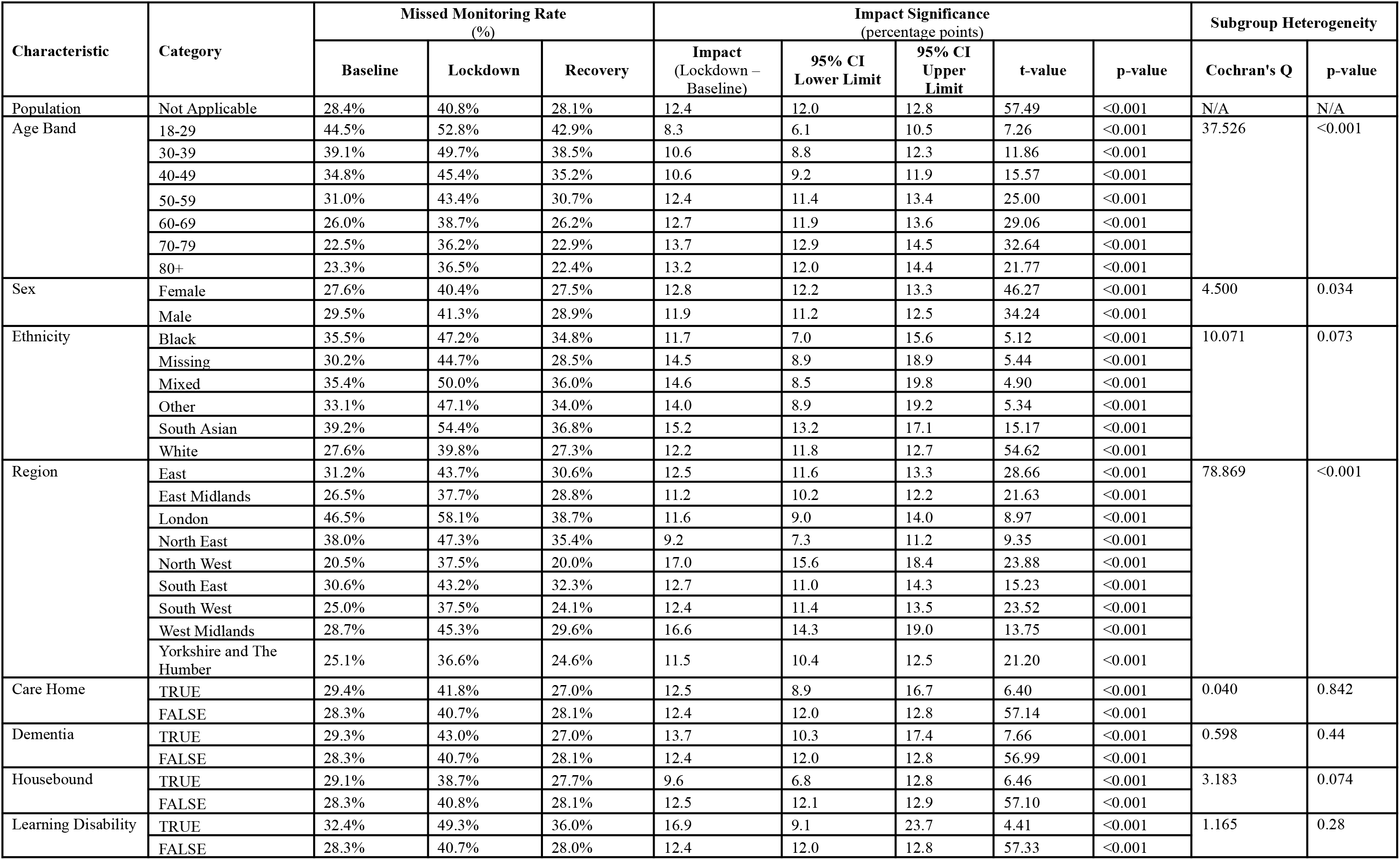
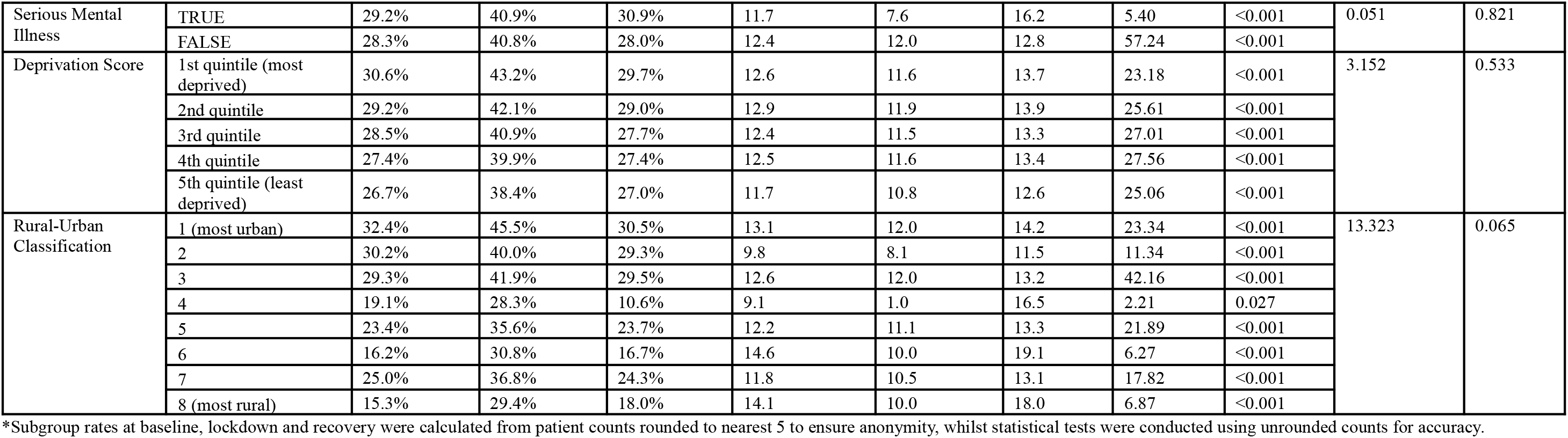
DMARD missed monitoring rates* for the population broken down into demographic and clinical characteristics at baseline (Dec19 to Feb20), lockdown (Mar20 to May20) and recovery (May22 to Jul22).

| Characteristic | Category | Missed Monitoring Rate (%) |  |  | Impact Significance (percentage points) |  |  |  |  | Subgroup Heterogeneity |  |
| --- | --- | --- | --- | --- | --- | --- | --- | --- | --- | --- | --- |
|  |  | Baseline | Lockdown | Recovery | Impact (Lockdown – Baseline) | 95% CI Lower Limit | 95% CI Upper Limit | t-value | p-value | Cochran's Q | p-value |
| Population | Not Applicable | 28.4% | 40.8% | 28.1% | 12.4 | 12.0 | 12.8 | 57.49 | <0.001 | N/A | N/A |
| Age Band | 18-29 | 44.5% | 52.8% | 42.9% | 8.3 | 6.1 | 10.5 | 7.26 | <0.001 | 37.526 | <0.001 |
|  | 30-39 | 39.1% | 49.7% | 38.5% | 10.6 | 8.8 | 12.3 | 11.86 | <0.001 |  |  |
|  | 40-49 | 34.8% | 45.4% | 35.2% | 10.6 | 9.2 | 11.9 | 15.57 | <0.001 |  |  |
|  | 50-59 | 31.0% | 43.4% | 30.7% | 12.4 | 11.4 | 13.4 | 25.00 | <0.001 |  |  |
|  | 60-69 | 26.0% | 38.7% | 26.2% | 12.7 | 11.9 | 13.6 | 29.06 | <0.001 |  |  |
|  | 70-79 | 22.5% | 36.2% | 22.9% | 13.7 | 12.9 | 14.5 | 32.64 | <0.001 |  |  |
|  | 80+ | 23.3% | 36.5% | 22.4% | 13.2 | 12.0 | 14.4 | 21.77 | <0.001 |  |  |
| Sex | Female | 27.6% | 40.4% | 27.5% | 12.8 | 12.2 | 13.3 | 46.27 | <0.001 | 4.500 | 0.034 |
|  | Male | 29.5% | 41.3% | 28.9% | 11.9 | 11.2 | 12.5 | 34.24 | <0.001 |  |  |
| Ethnicity | Black | 35.5% | 47.2% | 34.8% | 11.7 | 7.0 | 15.6 | 5.12 | <0.001 | 10.071 | 0.073 |
|  | Missing | 30.2% | 44.7% | 28.5% | 14.5 | 8.9 | 18.9 | 5.44 | <0.001 |  |  |
|  | Mixed | 35.4% | 50.0% | 36.0% | 14.6 | 8.5 | 19.8 | 4.90 | <0.001 |  |  |
|  | Other | 33.1% | 47.1% | 34.0% | 14.0 | 8.9 | 19.2 | 5.34 | <0.001 |  |  |
|  | South Asian | 39.2% | 54.4% | 36.8% | 15.2 | 13.2 | 17.1 | 15.17 | <0.001 |  |  |
|  | White | 27.6% | 39.8% | 27.3% | 12.2 | 11.8 | 12.7 | 54.62 | <0.001 |  |  |
| Region | East | 31.2% | 43.7% | 30.6% | 12.5 | 11.6 | 13.3 | 28.66 | <0.001 | 78.869 | <0.001 |
|  | East Midlands | 26.5% | 37.7% | 28.8% | 11.2 | 10.2 | 12.2 | 21.63 | <0.001 |  |  |
|  | London | 46.5% | 58.1% | 38.7% | 11.6 | 9.0 | 14.0 | 8.97 | <0.001 |  |  |
|  | North East | 38.0% | 47.3% | 35.4% | 9.2 | 7.3 | 11.2 | 9.35 | <0.001 |  |  |
|  | North West | 20.5% | 37.5% | 20.0% | 17.0 | 15.6 | 18.4 | 23.88 | <0.001 |  |  |
|  | South East | 30.6% | 43.2% | 32.3% | 12.7 | 11.0 | 14.3 | 15.23 | <0.001 |  |  |
|  | South West | 25.0% | 37.5% | 24.1% | 12.4 | 11.4 | 13.5 | 23.52 | <0.001 |  |  |
|  | West Midlands | 28.7% | 45.3% | 29.6% | 16.6 | 14.3 | 19.0 | 13.75 | <0.001 |  |  |
|  | Yorkshire and The Humber | 25.1% | 36.6% | 24.6% | 11.5 | 10.4 | 12.5 | 21.20 | <0.001 |  |  |
| Care Home | TRUE | 29.4% | 41.8% | 27.0% | 12.5 | 8.9 | 16.7 | 6.40 | <0.001 | 0.040 | 0.842 |
|  | FALSE | 28.3% | 40.7% | 28.1% | 12.4 | 12.0 | 12.8 | 57.14 | <0.001 |  |  |
| Dementia | TRUE | 29.3% | 43.0% | 27.0% | 13.7 | 10.3 | 17.4 | 7.66 | <0.001 | 0.598 | 0.44 |
|  | FALSE | 28.3% | 40.7% | 28.1% | 12.4 | 12.0 | 12.8 | 56.99 | <0.001 |  |  |
| Housebound | TRUE | 29.1% | 38.7% | 27.7% | 9.6 | 6.8 | 12.8 | 6.46 | <0.001 | 3.183 | 0.074 |
|  | FALSE | 28.3% | 40.8% | 28.1% | 12.5 | 12.1 | 12.9 | 57.10 | <0.001 |  |  |
| Learning Disability | TRUE | 32.4% | 49.3% | 36.0% | 16.9 | 9.1 | 23.7 | 4.41 | <0.001 | 1.165 | 0.28 |
|  | FALSE | 28.3% | 40.7% | 28.0% | 12.4 | 12.0 | 12.8 | 57.33 | <0.001 |  |  |
| Serious Mental Illness | TRUE | 29.2% | 40.9% | 30.9% | 11.7 | 7.6 | 16.2 | 5.40 | <0.001 | 0.051 | 0.821 |
|  | FALSE | 28.3% | 40.8% | 28.0% | 12.4 | 12.0 | 12.8 | 57.24 | <0.001 |  |  |
| Deprivation Score | 1st quintile (most deprived) | 30.6% | 43.2% | 29.7% | 12.6 | 11.6 | 13.7 | 23.18 | <0.001 | 3.152 | 0.533 |
|  | 2nd quintile | 29.2% | 42.1% | 29.0% | 12.9 | 11.9 | 13.9 | 25.61 | <0.001 |  |  |
|  | 3rd quintile | 28.5% | 40.9% | 27.7% | 12.4 | 11.5 | 13.3 | 27.01 | <0.001 |  |  |
|  | 4th quintile | 27.4% | 39.9% | 27.4% | 12.5 | 11.6 | 13.4 | 27.56 | <0.001 |  |  |
|  | 5th quintile (least deprived) | 26.7% | 38.4% | 27.0% | 11.7 | 10.8 | 12.6 | 25.06 | <0.001 |  |  |
| Rural-Urban Classification | 1 (most urban) | 32.4% | 45.5% | 30.5% | 13.1 | 12.0 | 14.2 | 23.34 | <0.001 | 13.323 | 0.065 |
|  | 2 | 30.2% | 40.0% | 29.3% | 9.8 | 8.1 | 11.5 | 11.34 | <0.001 |  |  |
|  | 3 | 29.3% | 41.9% | 29.5% | 12.6 | 12.0 | 13.2 | 42.16 | <0.001 |  |  |
|  | 4 | 19.1% | 28.3% | 10.6% | 9.1 | 1.0 | 16.5 | 2.21 | 0.027 |  |  |
|  | 5 | 23.4% | 35.6% | 23.7% | 12.2 | 11.1 | 13.3 | 21.89 | <0.001 |  |  |
|  | 6 | 16.2% | 30.8% | 16.7% | 14.6 | 10.0 | 19.1 | 6.27 | <0.001 |  |  |
|  | 7 | 25.0% | 36.8% | 24.3% | 11.8 | 10.5 | 13.1 | 17.82 | <0.001 |  |  |
|  | 8 (most rural) | 15.3% | 29.4% | 18.0% | 14.1 | 10.0 | 18.0 | 6.87 | <0.001 |  |  |

\*Subgroup rates at baseline, lockdown and recovery were calculated from patient counts rounded to nearest 5 to ensure anonymity, whilst statistical tests were conducted using unrounded counts for accuracy.

## Supplementary Figures

**Figure S1.**
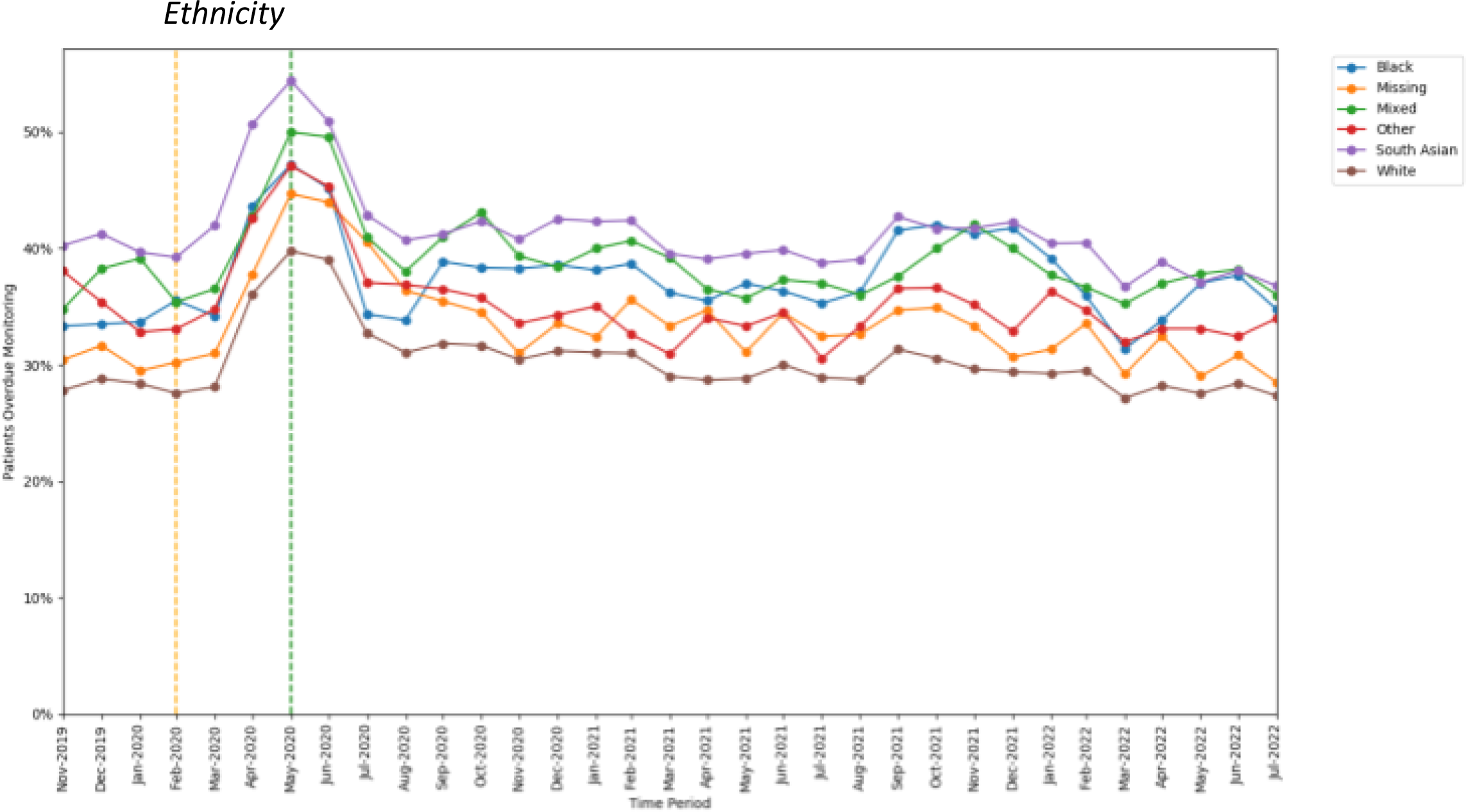
Proportions of patients overdue DMARD monitoring between November 2019 to July 2022, broken down by ethnicity. The baseline period before lockdown is shown as an orange dashed vertical line. The monitoring window, measured as 3 months from the onset of the March 2020 COVID-19 lockdown, is shown as a green dashed vertical line.

**Figure S2.**
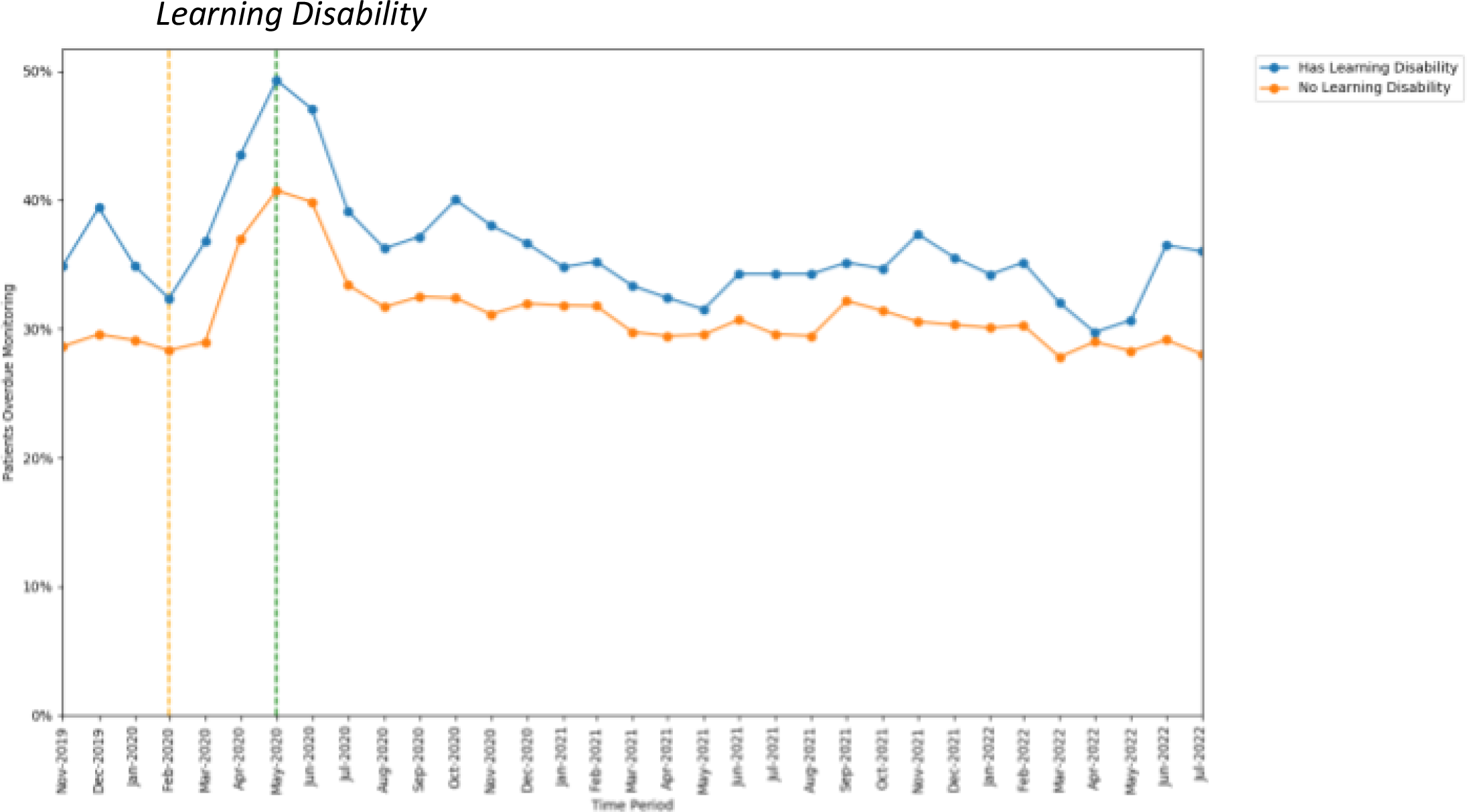
Proportions of patients overdue DMARD monitoring between November 2019 to July 2022, broken down by whether a learning disability diagnosis was coded. The baseline period before lockdown is shown asan orange dashed vertical line. The monitoring window, measured as 3 months from the onset of the March 2020 COVID-19 lockdown, is shown as a green dashed vertical line.

**Figure S3.**
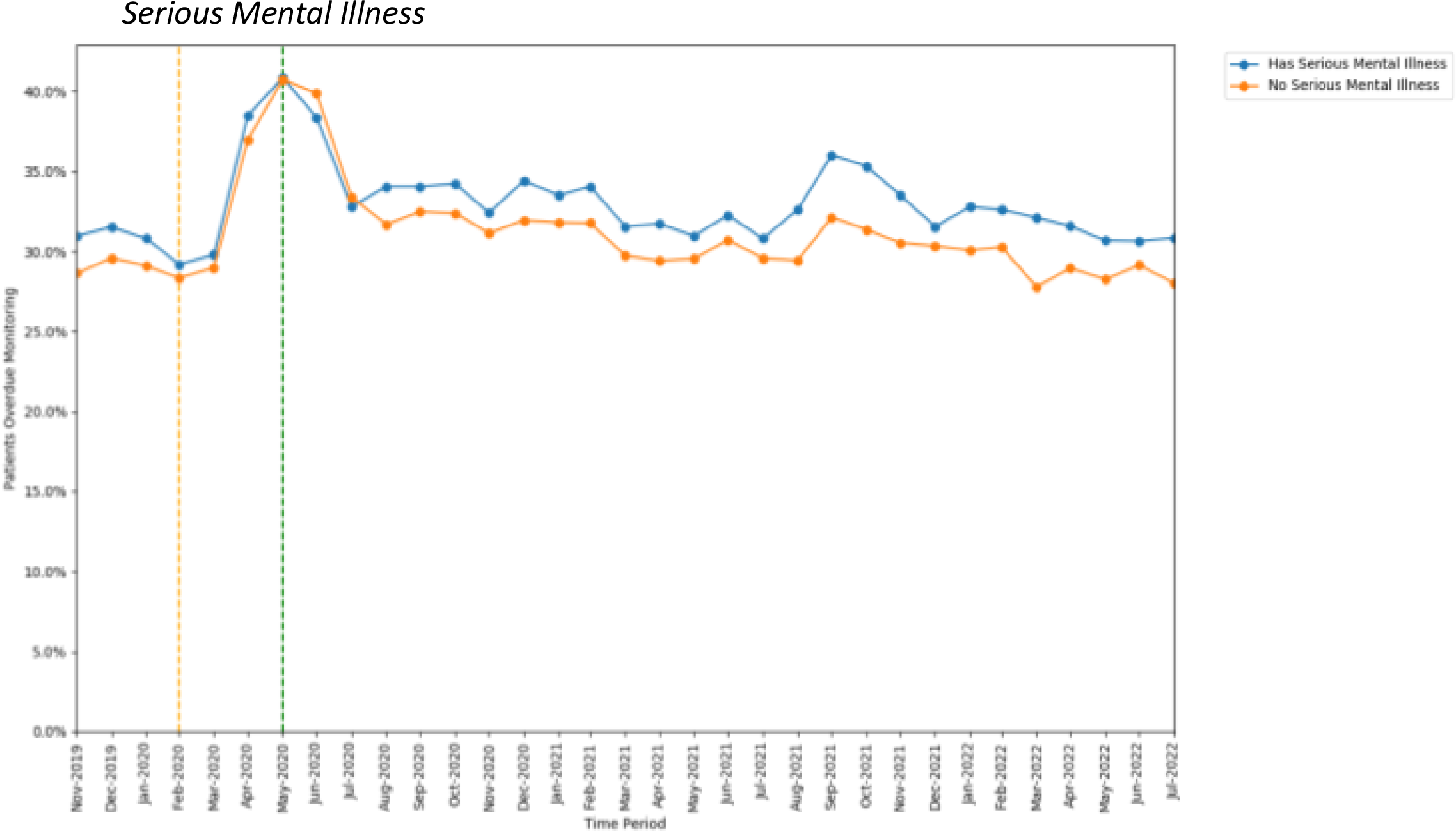
Proportions of patients overdue DMARD monitoring between November 2019 to July 2022, broken down by whether a serious mental illness diagnosis was coded. The baseline period before lockdown is shown asan orange dashed vertical line. The monitoring window, measured as 3 months from the onset of the Mareh 2020 COVID-19 lockdown, is shown as a green dashed vertical line.

**Figure S4.**
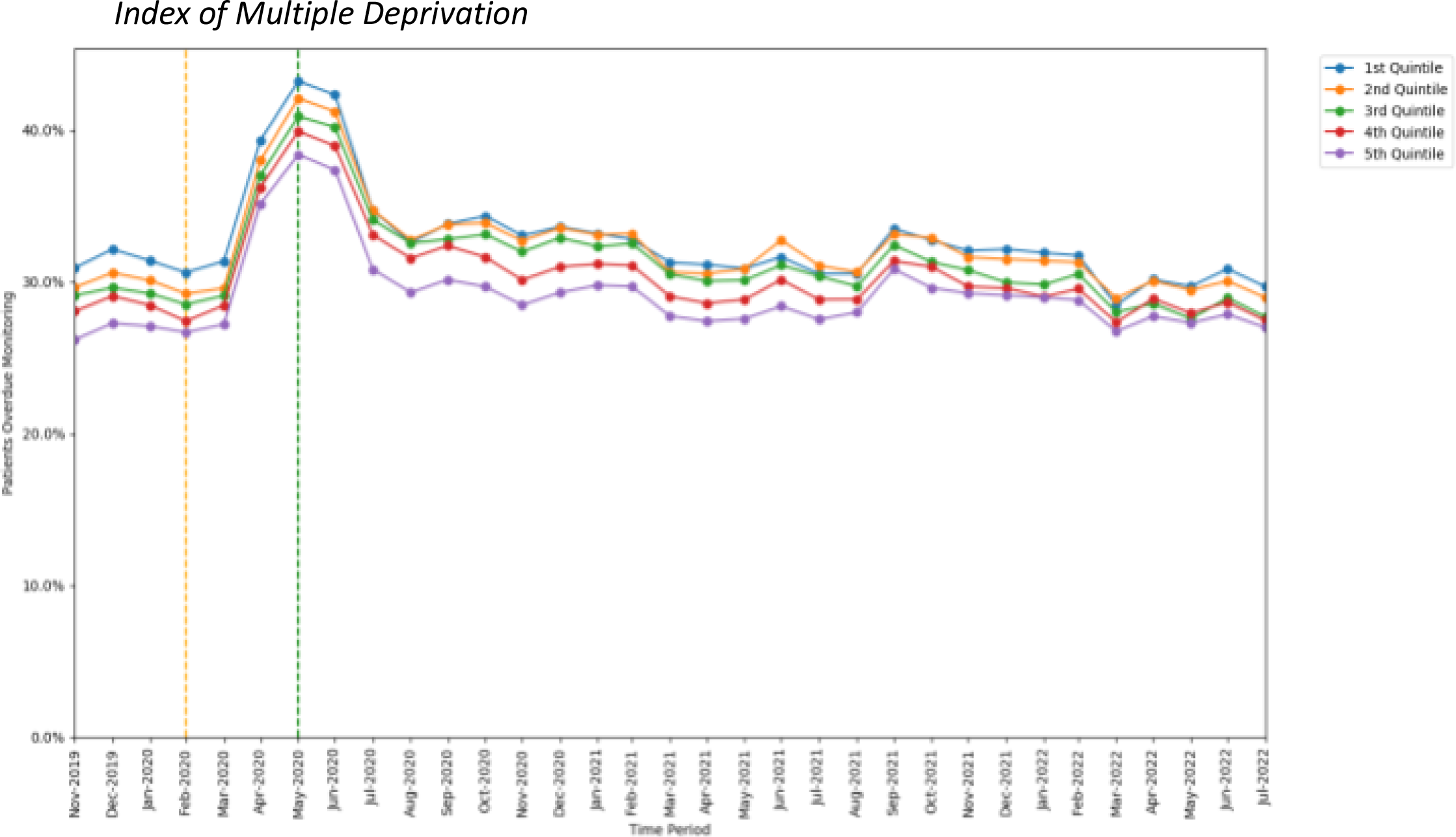
Proportions of patients overdue DMARD monitoring between November 2019 to July 2022, broken down by Index of Multiple Deprivation quintile. The baseline period before lockdown is shown as an orange dashed vertical line. The monitoring window, measured as 3 months from the onset of the March 2020 COVID-19 lockdown, is shown as a green dashed vertical line.

**Figure S5.**
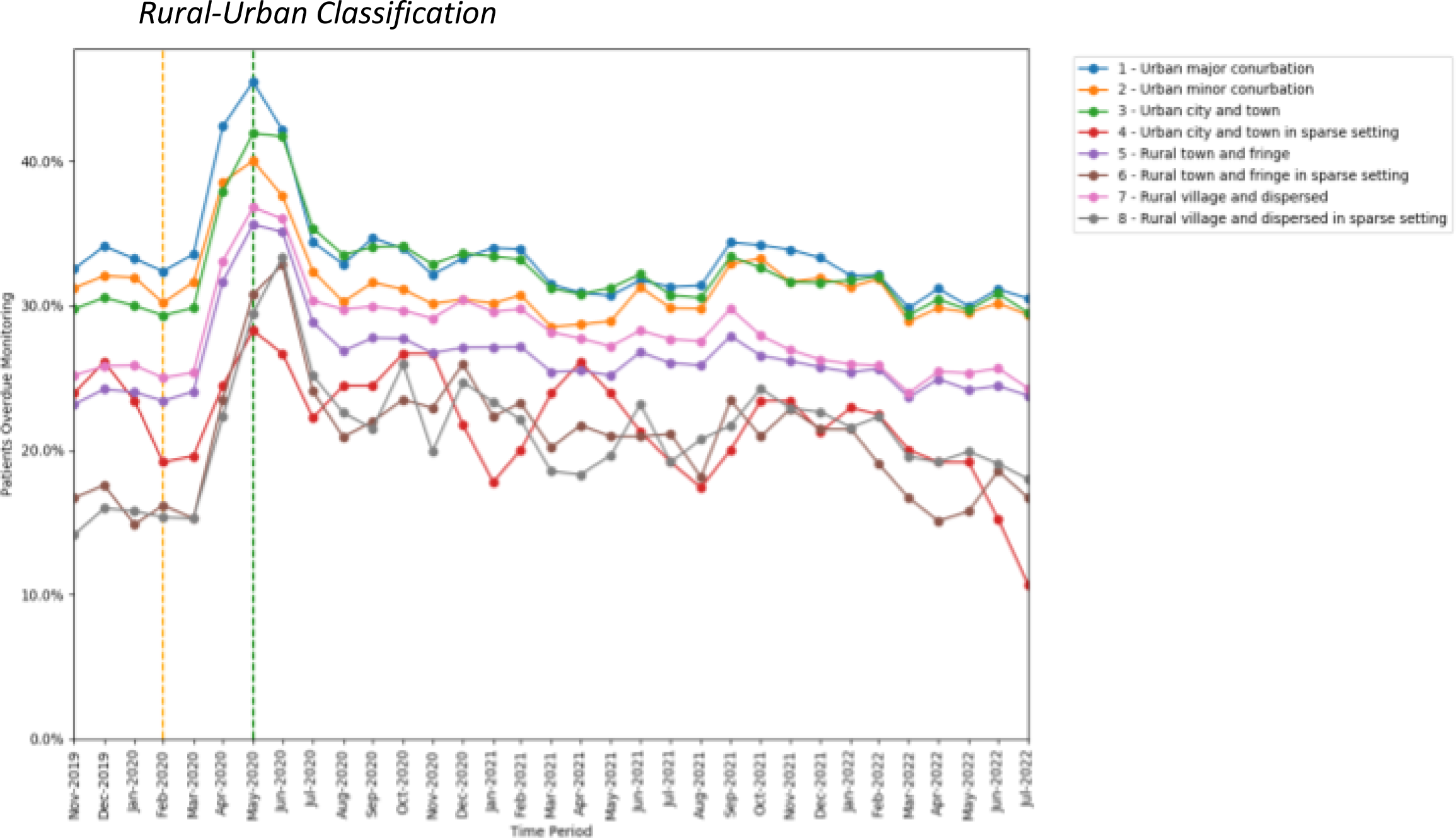
Proportions of patients overdue DMARD monitoring between November 2019 to July 2022, broken down by rural-urban score band. The baseline period hcfore lockdown is shown as an orange dashed vertical line. The monitoring window, measured as 3 months from the onset of the March 2020 COVID-19 lockdown, is shown as a green dashed vertical line.

**Figure S6.**
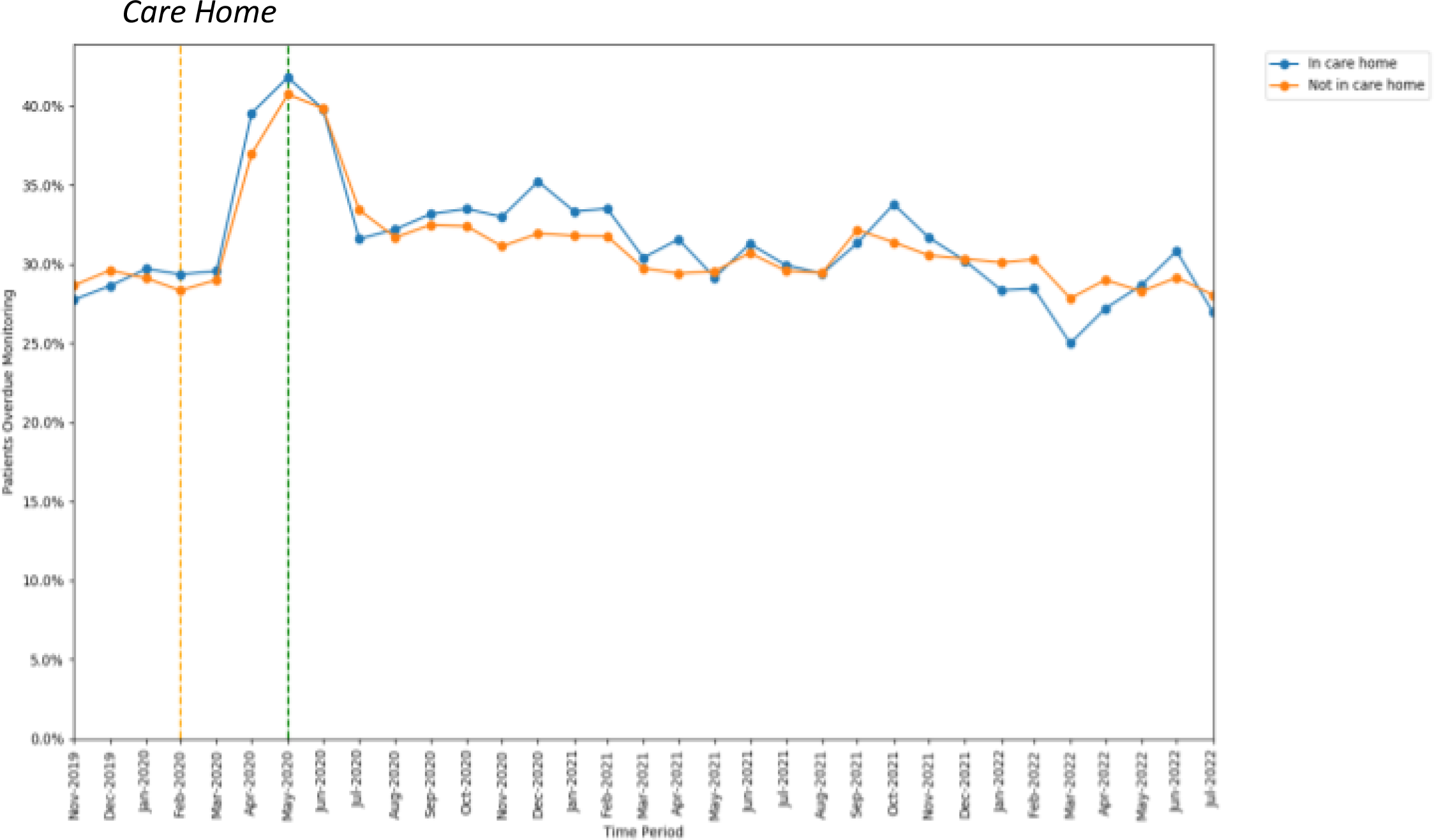
Proportions of patients overdue DMARD monitoring between November 2019 to July 2022, broken down by whether patients were coded as residing in a care home. The baseline period before lockdown is shown as an orange dashed vertical line, The monitoring window, measured as 3 months from the onset of the March 2020 COVID-19 lockdown, is shown as a green dashed vertical line.

**Figure S7.**
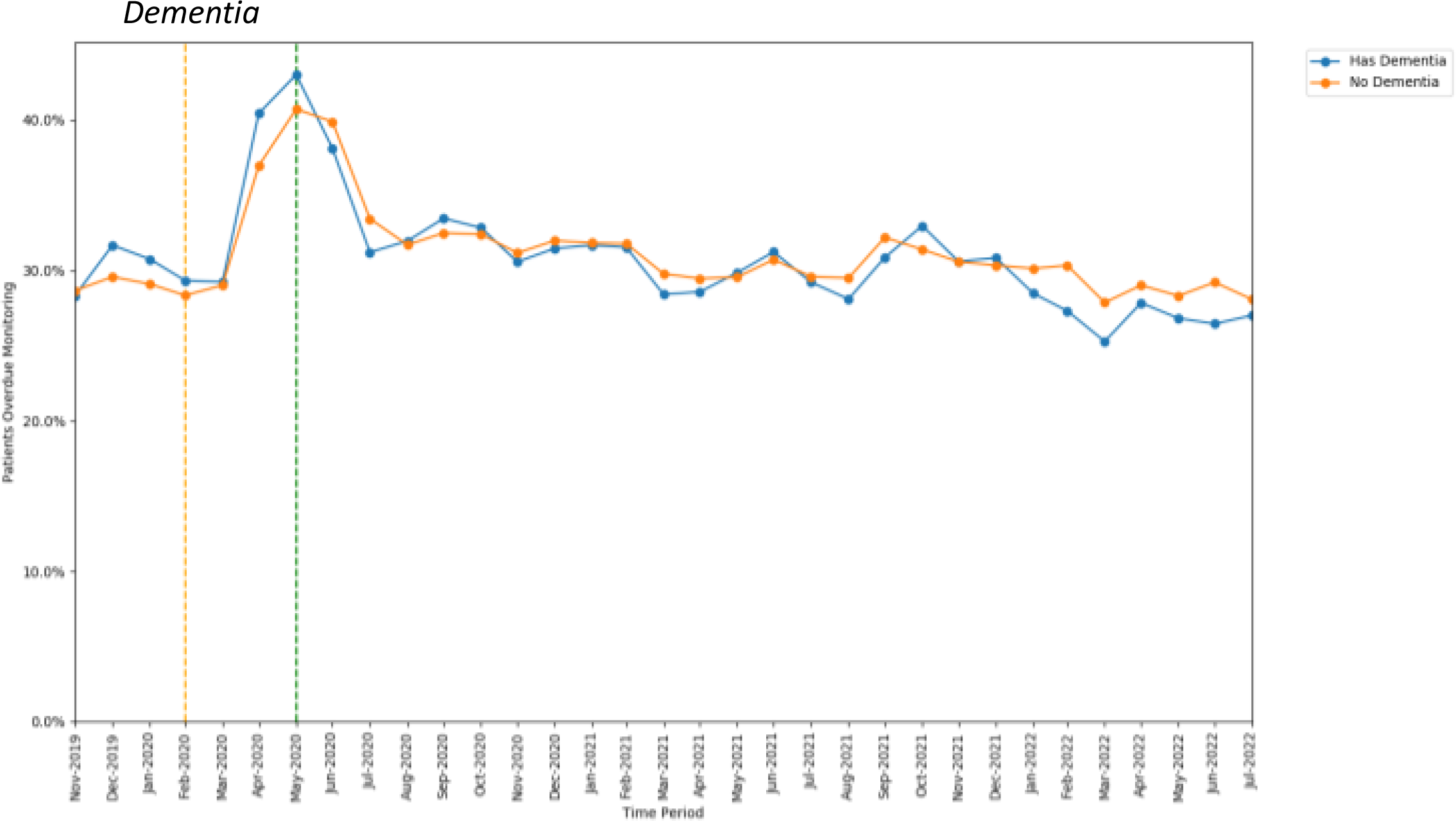
Proportions of patients overdue DMARD monitoring between November 2019 to July 2022, broken down by whether a dementia diagnosis was coded. The baseline period before lockdown is shown as an orange dashed vertical line. The monitoring window, measured as 3 months from the onset of the March 2D20 COVID-19 lockdown, is shown as a green dashed vertical line.

**Figure S8.**
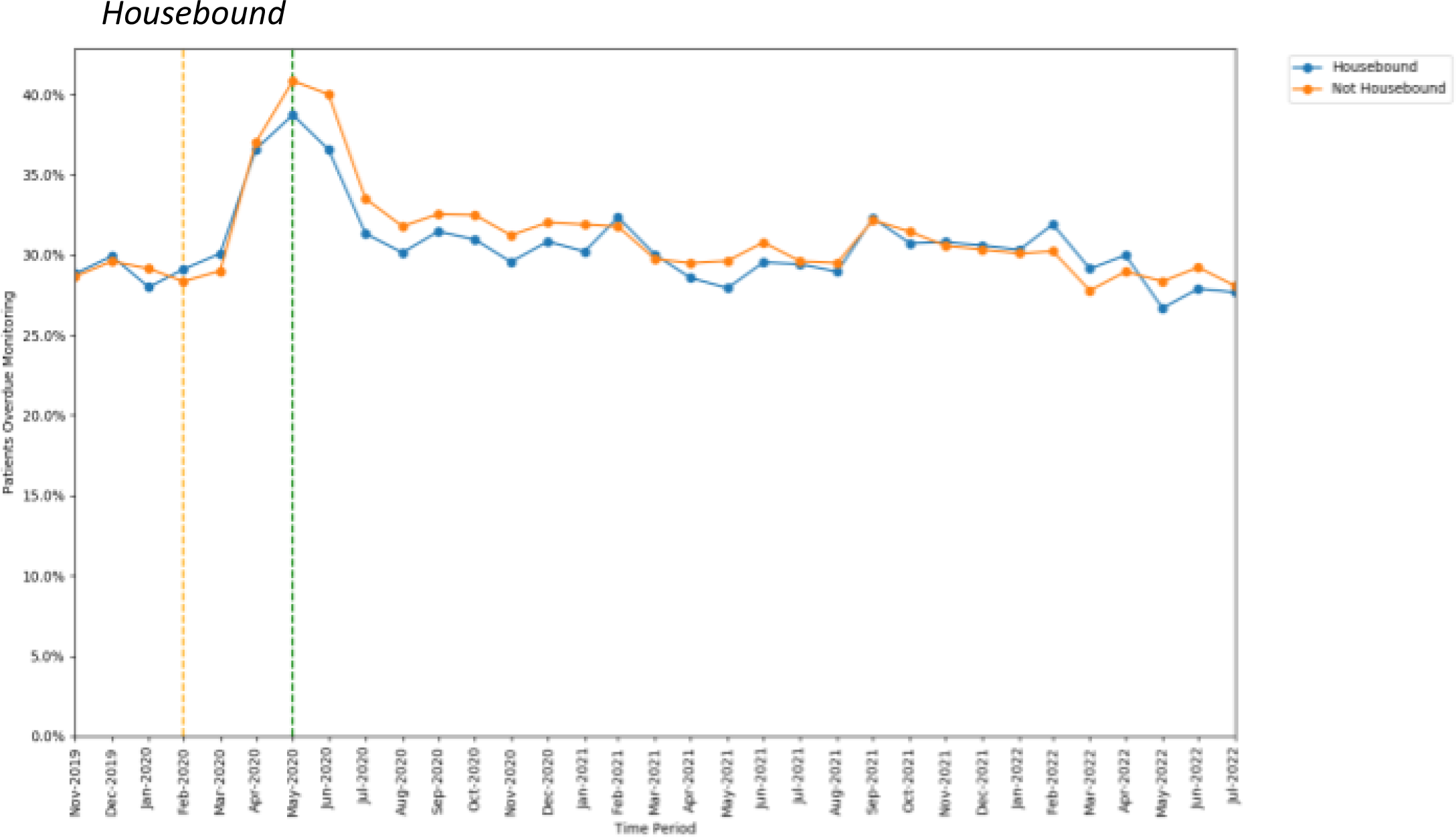
Proportions of patients overdue DMARD monitoring between November 2019 to July 2022, broken down by whether patients were coded as housebound. The baseline period before lockdown is shown as an orange dashed vertical line. The monitoring window, measured as 3 months from the onset of the Mareh 2020 COVID-19 Iockdown, is shawn as a green dashed vertical line.

